# A modular approach to forecasting COVID-19 hospital bed occupancy

**DOI:** 10.1101/2024.10.13.24314968

**Authors:** Ruarai J Tobin, Camelia R Walker, Robert Moss, James M McCaw, David J Price, Freya M Shearer

## Abstract

Monitoring the number of COVID-19 patients in hospital beds was a critical component of Australia’s real-time surveillance strategy for the disease. From 2021–2023, we produced short-term forecasts of bed occupancy to support public health decision making. In this work, we present a model for forecasting the number of ward and intensive care unit (ICU) beds occupied by COVID-19 cases. The model simulates the stochastic progression of COVID-19 patients through the hospital system and is fit to reported occupancy counts using an approximate Bayesian method. We do not directly model infection dynamics — instead taking independently produced forecasts of case incidence as an input — enabling the independent development of our model from that of the underlying case forecast(s). We evaluate the performance of 21-day forecasts of ward and ICU occupancy across Australia’s eight states and territories between March and September 2022, when major waves of the Omicron variant of SARS-CoV-2 were occurring throughout the country. Forecasts were on average biased downwards immediately prior to epidemic peaks and biased upwards post-peak. Forecast performance was best in jurisdictions with the largest population sizes. Our forecasts of COVID-19 hospital burden were reported weekly to national decision-making committees to support Australia’s public health response.

## Introduction

Throughout 2020–2022, SARS-CoV-2 induced large epidemic waves of infection internationally, with a considerable proportion of these infections requiring medical care. During peak epidemic periods, the demand for hospital beds overwhelmed the capacity of healthcare systems in many settings [23, 15, 13]. The number of beds occupied by COVID-19 cases depends upon the number of new patients admitted and the length of stay of these patients — with both quantities being products of the severity of disease and of clinical practice. Forecasts of hospital occupancy can provide public health decision makers with intelligence to support decision-making.

Australia’s early COVID-19 experience differed from most other countries, with only a small proportion of the population having been infected prior to the widespread uptake of vaccination; by December 2021, over 80% of adults had been vaccinated and less than 2% of adults had been recorded as infected amidst intensive public health measures [14, 43]. The Omicron variant of SARS-CoV-2 emerged in November 2021, with the Omicron BA.1 lineage inducing major waves of infection across Australia and resulting in at least 17% of the population having been infected by March 2022 [27]. We limit our study to the period between March and September 2022, which was defined by two major waves of infection: a wave induced by the Omicron BA.2 lineage, which peaked in March–April 2022 [10]; and a wave induced by the Omicron BA.4 and BA.5 lineages, which peaked in late July 2022 [9].

In this work, we describe a model for producing short-term (21-day) forecasts of hospital occupancy. We chose daily bed occupancy as a forecast target — rather than daily admissions — as occupancy more closely relates to the overall capacity of the hospital system. Furthermore, such bed occupancy counts had been collected and publicly reported for each state and territory of Australia on a daily basis since the early stages of the pandemic [32]. Our forecasting model takes as input an independently produced forecast of daily case incidence, with this incidence then transformed into ward and ICU occupancy counts through a stochastic compartmental model, with the probabilities of hospitalisation and of ICU admission informed by near-real-time data. The duration of time spent in each compartment is informed by censoring-adjusted estimates of patient length of stay, with estimation of these quantities described in our previous work [46]. Simulation outputs are then fit to reported occupancy counts using an Approximate Bayesian Computation approach [45].

Under the specifications of the Australian National Disease Surveillance Plan for COVID-19 [7], forecasts from our model were reported to key national decision-making committees on a weekly basis as part of a national COVID-19 situational assessment program [42]. We examine the performance of the forecasts throughout the study period (March – September 2022), both qualitatively — using visual checks — and quantitatively — with the use of formal statistical metrics [22, 2, 17, 18]. We discuss how the performance of our occupancy forecasts changed with the epidemiological context and how it depended upon the performance of the input case forecasts.

## Methods

### Summary

We produced forecasts of the number of COVID-19 cases in hospital ward and ICU beds (i.e. the ward and ICU occupancies) on a weekly basis using a bespoke clinical forecasting pipeline (Figure 1). We simulated the pathways taken by COVID-19 cases through a hospital as flow through a compartmental model (Figure 2). Our clinical forecasting pipeline takes in three primary inputs: an ensemble case forecast, time-varying estimates of key epidemiological parameters (the age distribution of cases, the probability of hospital admission, and the probability of ICU admission), and estimates of patient length of stay. The model outputs are fit to reported occupancy counts across a seven-day window prior to the forecast start date using Approximate Bayesian Computation (ABC) [45]. We reported the resultant 21-day forecasted counts of ward and ICU occupancy to public health committees on a weekly basis.

**Figure 1:**
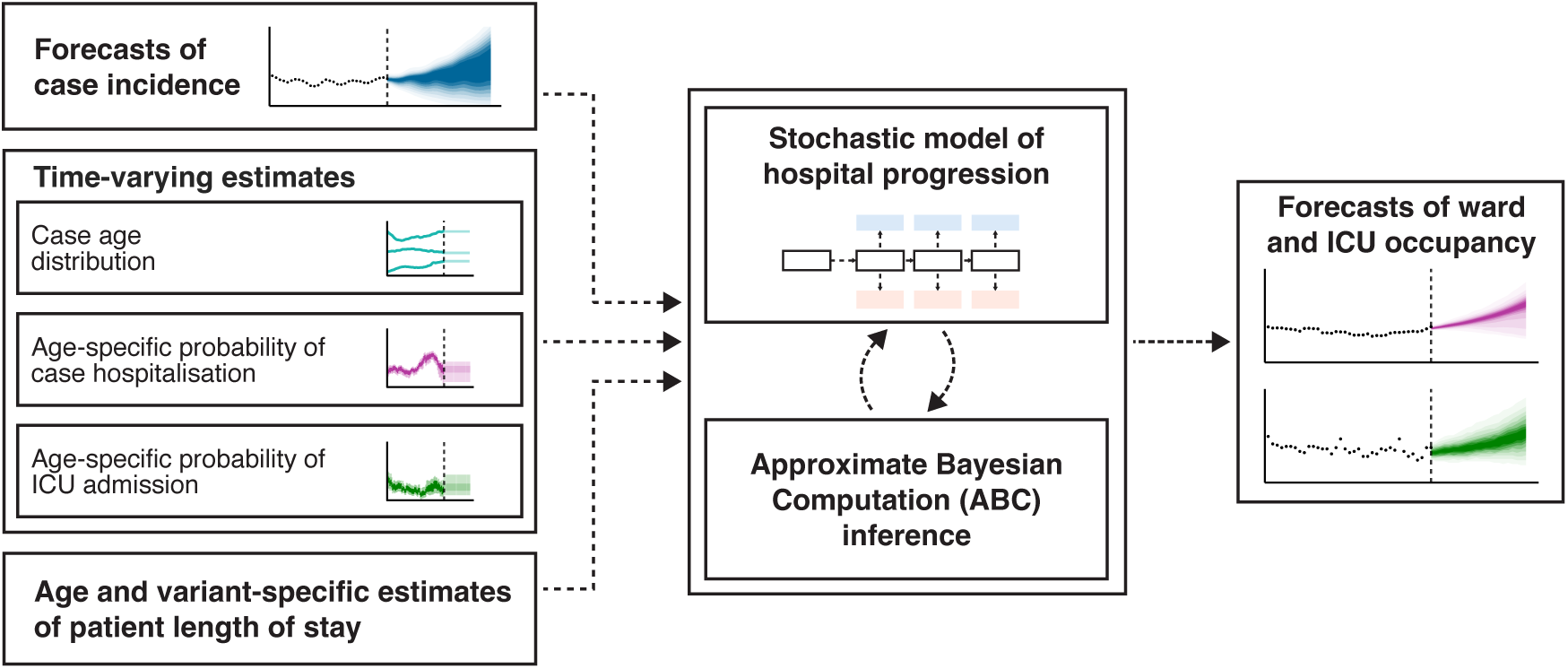
Overview of the clinical forecasting pipeline.

**Figure 2:**
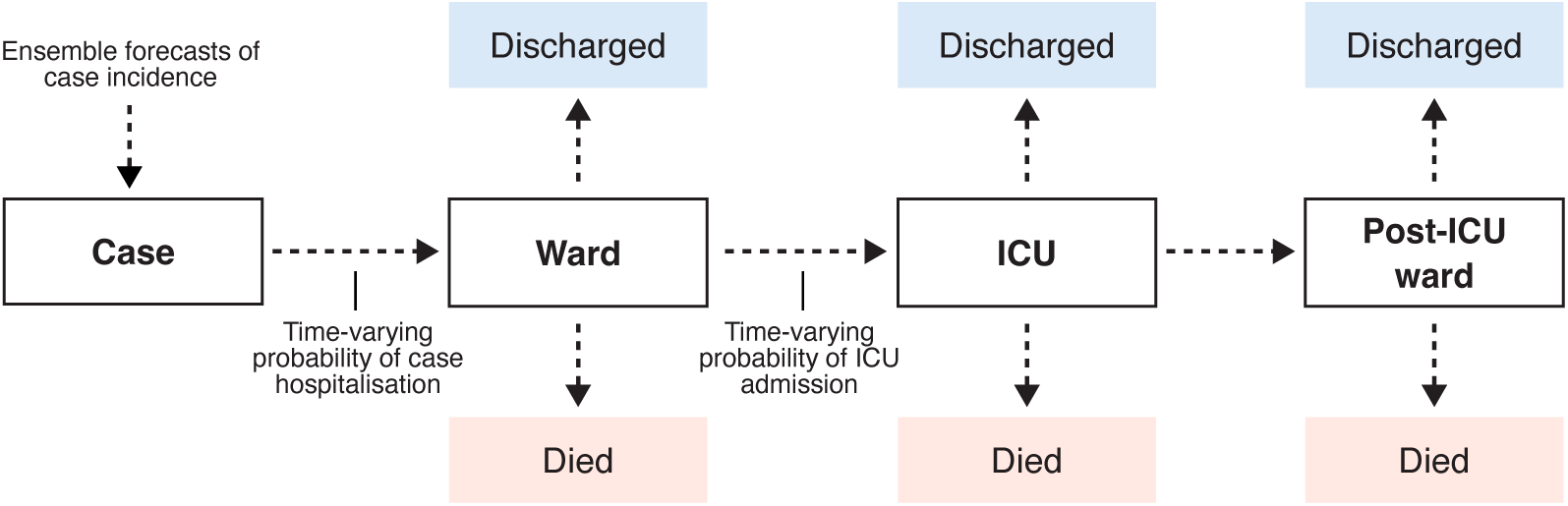
The compartmental model of COVID-19 clinical progression used to simulate the flow of COVID-19 patients through a hospital. The probability of transition between *Case* and *Ward* and between *Ward* and *ICU* were informed by time-varying age-specific estimates, all other probabilities were specified according to age-specific estimates from the multi-state length of stay analysis (estimation described elsewhere [46]). The number of occupied ward beds reported by the model is the sum of the individuals in the *Ward* and *Post-ICU ward* compartments, and the number of occupied ICU beds is the number of individuals in the *ICU* compartment.

### Compartmental pathways model

Our compartmental model simulates the progression of severe COVID-19 disease and corresponding pathways taken through a hospital (Figure 2). The design of this model was informed by COVID-19 clinical progression models previously developed for the Australian health system context [31, 39, 8]. In our model, new COVID-19 cases start in the *Case* compartment according to their date of symptom onset (inferred where not recorded). From this compartment, some fraction of cases are admitted to hospital, according to a (timevarying) probability of case hospitalisation. Hospitalisations start in the *Ward* compartment, from which a patient can then develop further severe disease and be admitted to *ICU*, according to a (time-varying) probability of ICU admission. Patients in the *ICU* compartment can then move to the *Post-ICU ward* compartment. In addition, across each of the *Ward*, *ICU*, and *Post-ICU ward* compartments, we assume patients have some probability of dying or being discharged. We count the number of occupied ward beds as the number of patients in the *Ward* and *Post-ICU ward* compartments, and the number of occupied ICU beds as the number of patients in the *ICU* compartment.

### Length of stay estimates

To simulate the flow of patients through the compartmental model, we need to specify distributional estimates of the duration of time they will spend within a compartment before a transition occurs (i.e. their length of stay), and the probabilities of each particular transition occurring (i.e. transition probabilities). We produced estimates of length of stay and transition probabilities using a multi-state survival analysis approach, with methods previously reported [46]. This survival analysis framework allowed us to (when necessary) produce estimates across our compartmental model in near-real-time while accounting for right-censoring, such that we could rapidly incorporate changes in length of stay or transition probabilities that may have occurred as a result of factors such as differences in the clinical severity of a new variant, changes in clinical practice, or vaccination [46]. We estimated length of stay and transition probabilities using hospital data from the state of New South Wales (see [46], Supp. Methods S4). The delay distribution for *Case* to *Ward* was informed by estimates (not described here) from the FluCAN sentinel hospital surveillance network study [6], as appropriate data were not available to estimate this delay in the New South Wales dataset. The transition probabilities from the multi-state survival model were used across all transitions in the compartmental model except for the *Case* to *Ward* and the *Ward* to *ICU* transitions. The transition probabilities for these two transitions were estimated as time-varying (described later) given their substantial impact upon the net occupancy counts. The length of stay and transition probability estimates were provided to the simulation model as bootstrapped samples of gamma distribution shape and scale parameters and multinomial probabilities of transition.

### Case incidence

In our compartmental model (Figure 2), cases of COVID-19 begin in the *Case* compartment. As such, we must inform the model with the number of new cases entering this compartment each day: we achieve this through use of a time series of historically reported case incidence concatenated with a trajectory of forecasted case incidence.

We received time series of historical case incidence indexed by date of symptom onset from an external model [20]. This external model performs imputation of symptom onset dates where they have not been recorded in the data, with the final time series being the count of cases with a (reported or imputed) onset date on each given date. Because this external model did not perform multiple imputation of the symptom onset date, we added noise to capture uncertainty in the case counts via sampling from a negative binomial distribution with a mean of the historical case count and a dispersion of *k =* 25.

Our method is agnostic to the case forecasting approach used as input, thus allowing us to couple it with any independently produced forecast of case incidence. Here we used outputs from an ensemble forecast of case incidence, which varied in model composition during the study period (methodologies and summary outputs for the ensemble forecast are publicly available [42]). A total of four different models were used at various stages: two mechanistic compartmental models, one mechanistic branching process model, and a non-mechanistic time series model (see [30, 19, 42] for details). Models within the ensemble received ongoing development across the study period in response to changes in our understanding of the epidemiology and biology of the virus [42].

### Estimation of time-varying parameters

We specified three parameters in the compartmental model of clinical progression as timevarying. For each forecast, we produced estimates stratified by age group *a* and varying with time *t* of: the probability of a case being within a certain age group, *p*_age_(*a*, *t*); the probability of a case being hospitalised, *p*_hosp_(*a*, *t*); and the probability of a hospitalised case being admitted to ICU, *p*_ICU_(*a*, *t*). These parameters were chosen to capture phenomena such as changes in case age distribution, changes in case ascertainment, differences in variant virulence and outbreaks of the disease within populations subgroups. We defined age groups as 10-year groups from age 0 to 80, followed by a final age group comprising individuals of age 80 and above (i.e. 0–9, 10–19, …, 80+).

The time-varying parameters were estimated using case data from the National Notifiable Disease Surveillance System (NNDSS), which collates information on COVID-19 cases across the eight state and territories of Australia. For each case in this dataset, we extracted the date of case notification, the recorded symptom onset date, the age of the case, and whether or not the case had been admitted to hospital or ICU. Where symptom onset date was not available, we assumed it to be one day prior to the date of notification (where this was the median delay observed in the data).

For each of the three time-varying parameters, we constructed estimates using a one-week moving-window average, with estimates for time *t* including all cases with a symptom onset date within the period (*t −*7, *t*]. We created 50 bootstrapped time series via sampling with replacement from the linelist data such that uncertainty in these estimates was propagated through the simulations. We calculated the first parameter *p*_age_(*a*, *t*), which defines the multinomial age distribution of cases over time, as the proportion of cases within each age group for an estimation window:

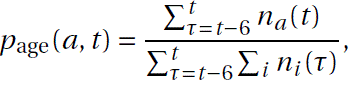

where *n_a_* (*τ*) is the number of cases in age group *a* with symptom onset at time *τ*. To calculate the probability of a case being hospitalised and the probability of a hospitalised case being admitted to ICU, we produced estimates with adjustment for right-truncation. Here, right-truncation was present as we used near-real-time epidemiological data and indexed our estimates by date of symptom onset. The most recent symptom onset dates in our estimates thus included cases that would eventually be (but had not yet been) hospitalised (and similarly for cases admitted to hospital, but not yet admitted to ICU). Had we not accounted for this right-truncation, we would have consistently underestimated the probabilities of hospitalisation and ICU admission for the most recent dates. We describe the maximum-likelihood estimation of the hospitalisation and ICU admission parameters in the supplementary materials (Supp. Methods S1).

If in a given reporting week we identified a jurisdiction as having unreliable data on hospitalised cases (most often, missing data on cases admitted to hospital or ICU due to data entry delays), we replaced the local estimates with estimates produced from pooled data across all other (reliable) jurisdictions. Changes made in this regard during the study period are listed in the supplementary material (Supp. Methods S5).

### Simulation and inference

To simulate a single trajectory of ward and ICU occupancy, we sampled: a time series of case incidence from the ensemble; a bootstrapped time series of the time-varying parameters; and a sample from the bootstrapped length of stay and transition probability estimates. Using these inputs, we performed simulations across the compartmental model (Figure 2) independently across each age group and then summed across all age groups to produce total ward and ICU counts for each day. The compartmental model simulates the pathways of patients through the hospital at the population-scale with an efficient agent-based approach; we provide details on this algorithm in the supplementary materials (Supp. Methods S2).

To ensure that trajectories simulated from the clinical pathways model aligned with reported occupancy counts, we introduce a simple rejection-sampling approximate Bayesian method, rejecting trajectories that did not match the true reported occupancy counts within a relative tolerance *ɛ* across a one-week calibration window. For each simulation with a simulated ward occupancy count *W*^^^ (*t*) and simulated ICU occupancy count *I*^^^(*t*), simulations were rejected where either:

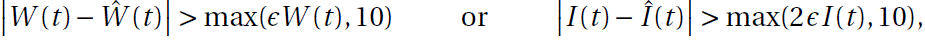

where *W* (*t*) and *I* (*t*) were the true reported occupancy counts for each date *t* in the fitting window, with these counts retrieved from the covid19data.com.au project [32]. We selected *ɛ* using a simple stepped threshold algorithm, initialising *ɛ* at a small value, and continued to sample simulations until 1,000 trajectories had been accepted by the model. If 1,000 trajectories were not accepted by the time that 100,000 simulations had been performed (i.e. 100 rejections per target number of output trajectories), we increased *ɛ* in sequence from [0.1, 0.2, 0.3, 0.5, 1, 10] and restarted the sampling procedure. This behaviour was chosen to achieve a good degree of predictive performance while ensuring that reporting deadlines were met (typically less than 24 hours from receipt of ensemble forecasts and relevant hospital data) even where the model was otherwise unlikely to capture hospital occupancy at tighter degrees of tolerance

We fit simulation outputs over a calibration window defined as the seven days following the start of the 28-day case forecast. This was chosen such that the most up-to-date occupancy data could be used in fitting (typically data as of, or a day prior to, the date clinical forecasts were produced). We could fit the clinical forecast over occupancy data points which were seven days in the future relative to the start of the case forecast for two reasons: the case forecasts were indexed by date of symptom onset and began at the date where a majority (>90%) of cases had experienced symptom onset, adding a delay of 2–3 days; and case forecasts were affected by reporting delays of 3–4 days (whereas occupancy data was not lagged). We did not fit over a larger window as the seven-day window was expected to be sufficient for our purposes and the computational requirements of model fitting would increase exponentially with a larger window. The forecasts we reported on a weekly basis and examine here are the model outputs across the 21 days following this seven-day fitting window.

We introduced two additional parameters to improve the ability of the model to fit to the reported occupancy counts. These parameters increased variance in the magnitude of the output ward and ICU occupancy count trajectories, reducing the probability of a substantial mismatch between these trajectories and the reported occupancy counts. The first parameter added was *H*, a modifier on the probability of hospitalisation acting linearly across logit-transformed values:

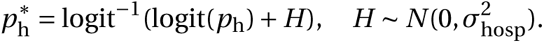

The second parameter added was *L*, which modified the shape of the length of stay distributions across the transitions out of *Case*, *Ward* and *Post-ICU Ward*, acting linearly across log-transformed values:

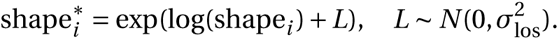

The values of *H* and *L* were sampled from normal distribution priors with means of zero and standard deviations of 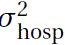 *=* 0.8 and 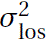 *=* 0.5 respectively. We specified these values to reduce the computational time required while ensuring the output model trajectories had good coverage over the reported occupancy counts. These parameters were changed for some jurisdictions during the study period, see the supplementary methods for details (Supp. Methods S5).

To illustrate the effect of the *H* and *L* parameters, we simulated model outputs for an example forecast with and without these parameters set to zero (Supp. Methods Figure 1A, B). This demonstrates that output trajectories without the effect of *H* and *L* may already align with the reported occupancy counts, but where this does not occur, they enable the recent reported occupancy counts to be well captured by the fitted model outputs (Supp. Methods Figure 1C).

### Performance evaluation

We consider the performance of our forecasts produced between March and September 2022. We produced plots for the visual assessment of forecast performance (Figure 3, Supp. Performance, Figures 5–20) which depict all forecasts across the study period with the same presentation of uncertainty as was used in official reporting of the forecasts (with pointwise credible intervals ranging from 20% through to 90% by steps of 10%, and reported occupancy counts overlaid).

**Figure 3:**
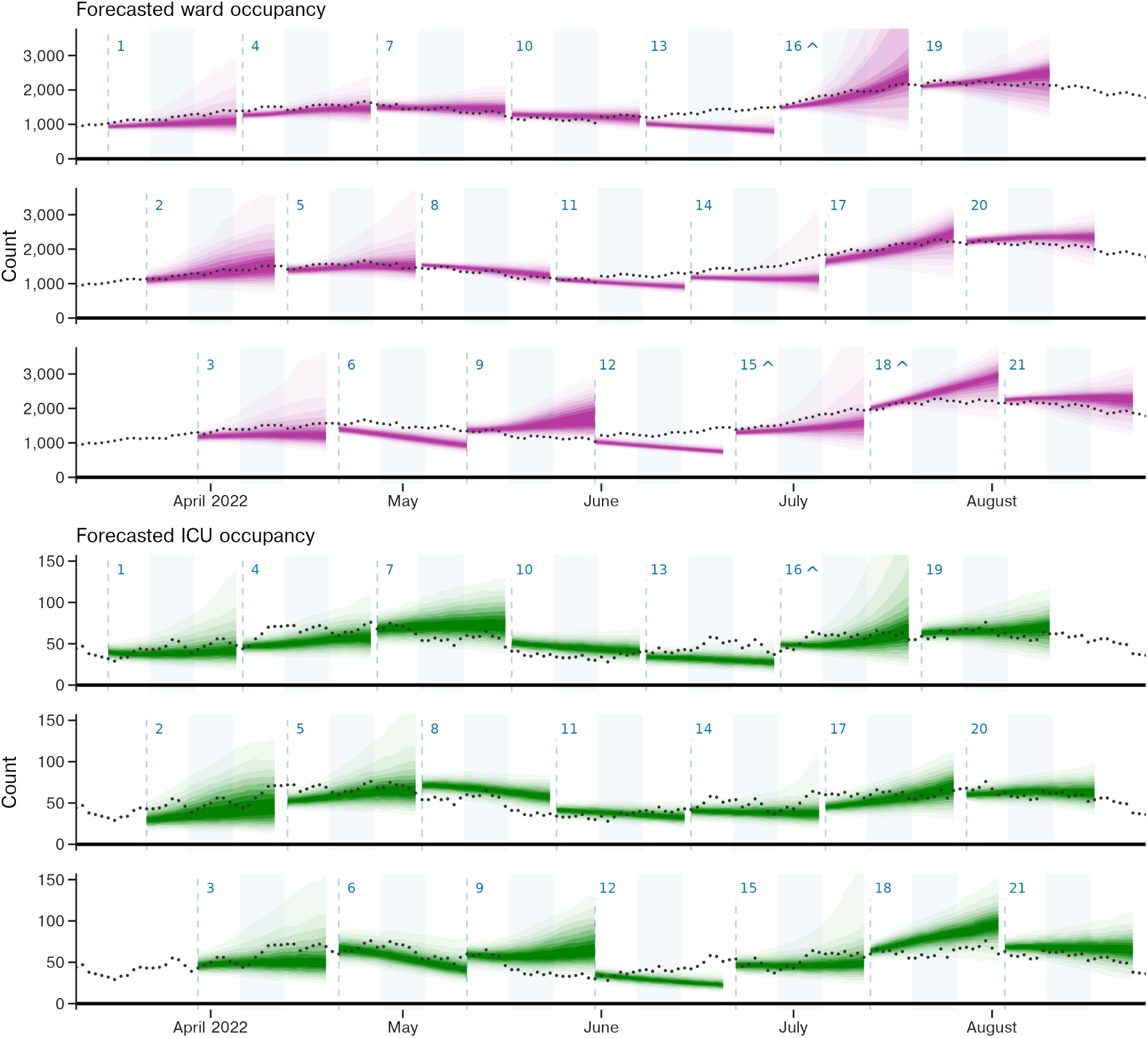
Forecasts of ward and ICU occupancy for the state of New South Wales produced between March and September 2022. Credible intervals from 20% through to 90% in 10% increments are displayed in progressively lighter shading. Reported occupancy counts are overlaid. As we produced our forecasts on a weekly basis and each forecast spans three weeks, forecasts are plotted interleaved across three rows; reported occupancy counts are repeated across each row. Forecast start dates are displayed as vertical dashed lines. Note that forecast start date was dependent upon that of the case forecast, and this varied slightly over time (see forecasts 5, 9, 12, and 19). The second week for each forecast (days 8–14) have background shaded in light blue. An identifier for each forecast, 1 through 21, is displayed above each forecast start and a ^ is displayed where the upper credible intervals of a forecast exceed the y-axis limits. Forecasts for other states and territories are provided in the supplementary materials.

To evaluate the overall performance of our forecasts, we calculated continuous ranked probability scores (CRPS) across log-transformed counts of occupancy. The CRPS measures the distributional accuracy of a set of forecasts against the eventual observations [22]. The CRPS is a proper scoring rule: in the limit, where a forecast reports the true probabilities of the underlying process, it will receive the greatest score. We calculated CRPS over log-transformed counts (specifically, 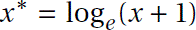) rather than over raw counts, as this has been argued to be more meaningful given the exponential nature of epidemic growth [2]. This transformation also allows us to interpret the resultant CRPS values as a relative error [2], enabling comparison of the forecast performance between different settings.

We calculated forecast bias to examine where the overall performance of our forecast was reduced due to consistent overprediction or underprediction [17] (Figure 5). Forecast bias (as opposed to, for example, estimator bias [48]) ranges between *−*1 and 1, with a bias greater than zero indicating overprediction and less than zero indicating underprediction. Bias values of approximately zero are ideal, indicating a forecast which overpredicts as often as it underpredicts (or vice versa).

We produced plots demonstrating the association between the performance of our ward and ICU occupancy forecasts and the underlying case forecasts used as input. Specifically, we compared the case forecast performance calculated using CRPS to the bias of the ward occupancy forecasts (Figure 6) and ICU occupancy forecasts (Supp. Performance, Figure 3), and the bias of the case forecasts to that of the ward occupancy forecasts (Supp. Performance, Figure 4). These values were calculated across the whole horizon of the respective forecasts; it should be noted that such comparisons are inherently limited due to the lag between onset of symptoms and admission to hospital, i.e. the performance of the case forecast at the 28 day horizon would be expected to be of lesser influence given these cases are less likely to be hospitalised within the time-frame of our simulation.

**Figure 4:**
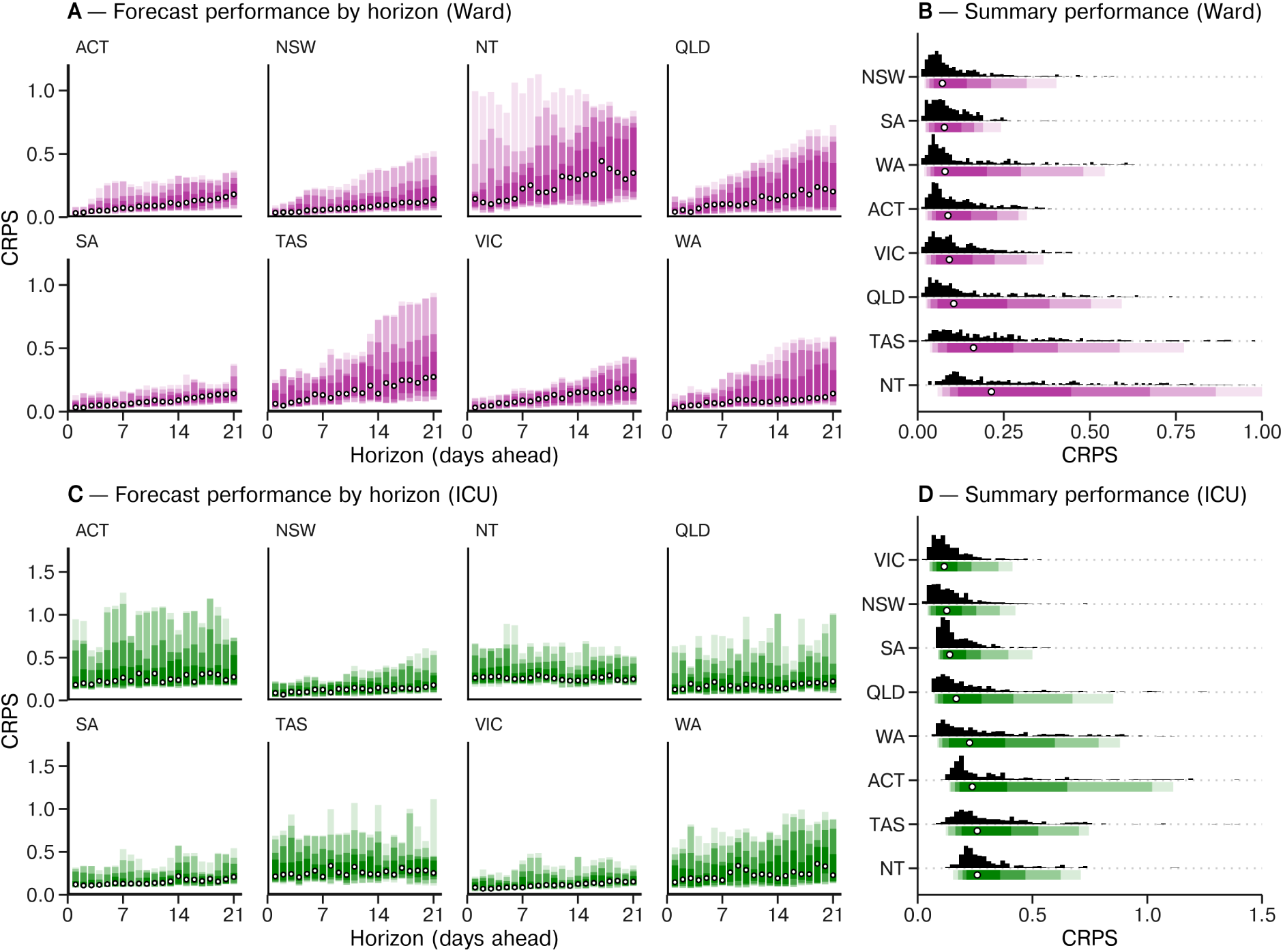
Summary performance of the ward and ICU forecasts across the study period (March to September 2022) measured using CRPS across log-transformed counts. ACT is the Australian Capital Territory, NSW is New South Wales, NT is the Northern Territory, QLD is Queensland, SA is South Australia, TAS is Tasmania, VIC is Victoria, and WA is Western Australia. **A**, **C**: Forecast performance for ward (top) and ICU (bottom) by forecast horizon. Median performance (in white) and intervals for 50%, 75%, 90%, 95% density (in purple or green) displayed. Note the differing x-axis scale across the ward and ICU forecast plots. **B**, **D**: Summary forecast performance for ward (top) and ICU (bottom) across all forecasting dates. Frequency for each state is displayed as a histogram (in black), and density underneath (in purple or green), with median overlaid (white and black points). States have been ordered according to median forecast performance. Note the differing x-axis scale across the ward and ICU forecast plots. Due to the limited x-axis scales, 14 points are omitted from the histogram for ward for the Northern Territory and 1 point for ICU for Tasmania.

**Figure 5:**
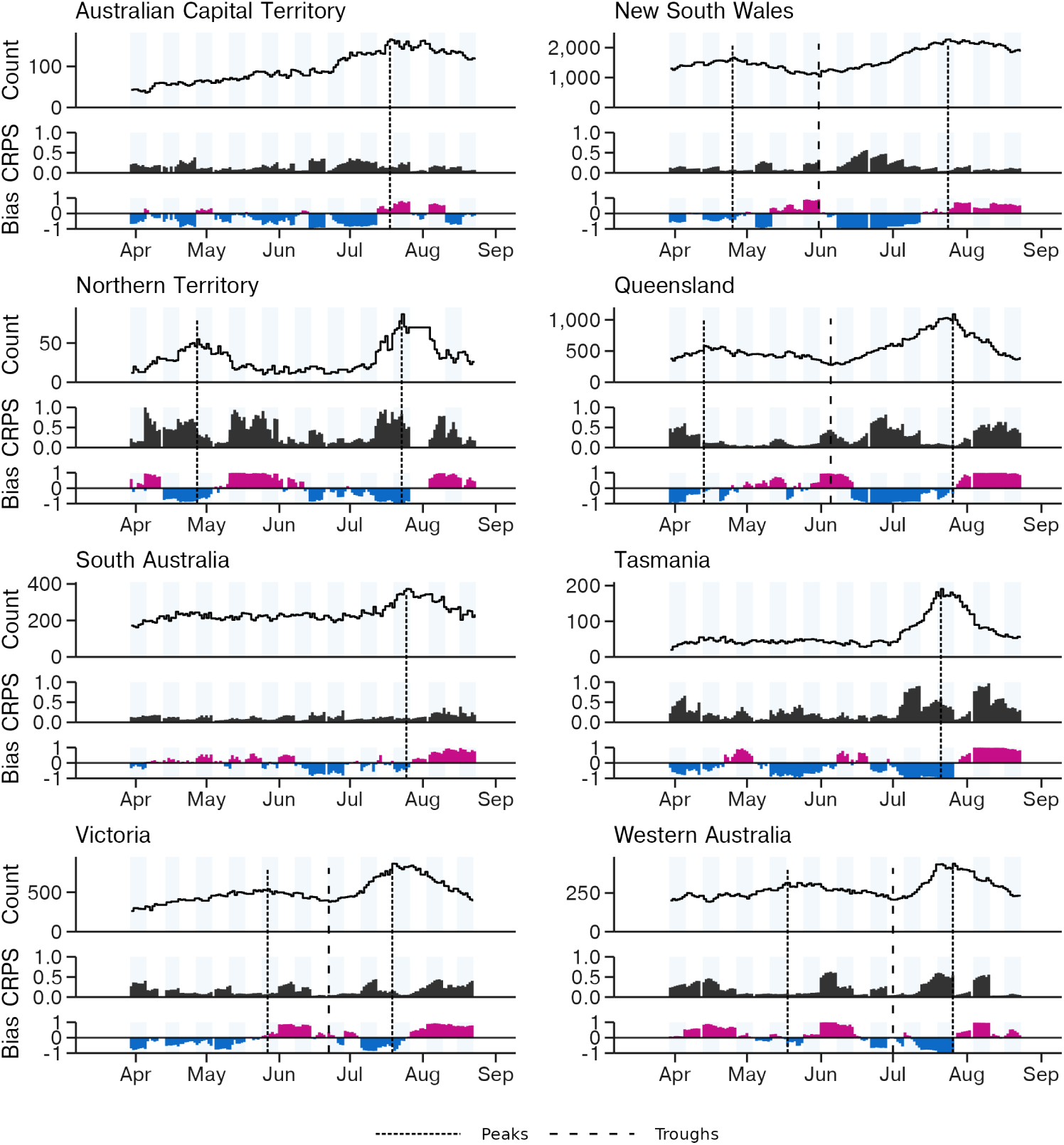
Three week (days 15–21) horizon performance of the ward forecasts for the forecasts produced between March and September 2022. Light blue shading indicates alternating forecast weeks. The true ward occupancy count is displayed at the top of each panel, with vertical dashed lines indicating dates of visually distinct peaks and troughs (dotted and dashed lines respectively) in the time series. The CRPS and bias of the forecast are displayed below, reflecting the performance of forecasted counts for that date within the 15–21 day forecast horizon. Upwards bias is displayed in magenta and downwards bias in blue. The CRPS is calculated over logtransformed counts. Optimal forecasting performance is achieved where these values are nearest to zero.

**Figure 6:**
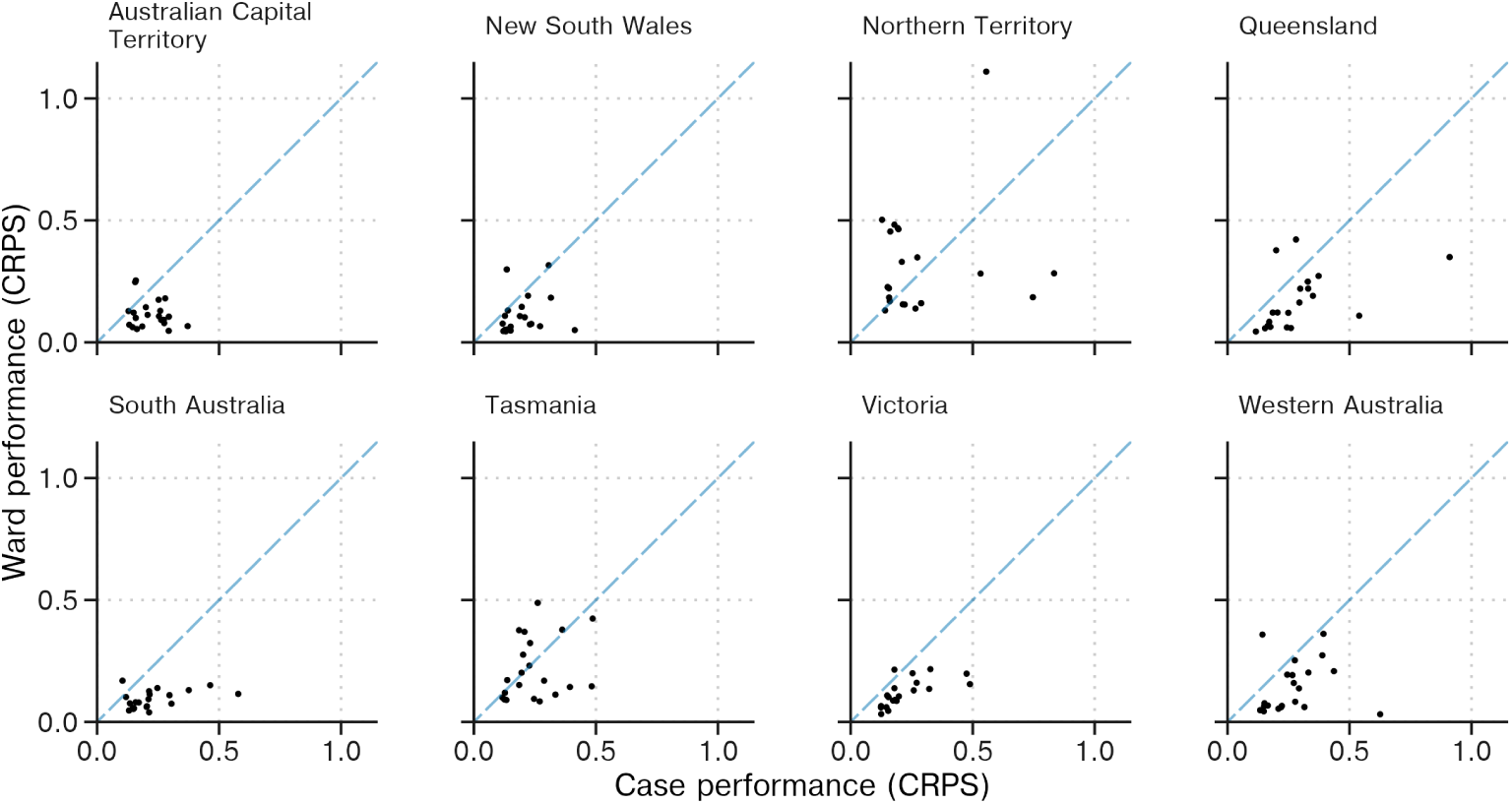
Performance of the ward occupancy forecasts (y-axis) in comparison to the corresponding ensemble case forecast used as input (x-axis), where performance is measured using CRPS over log-transformed counts. Each dot represents performance measured over a 28-day case incidence ensemble forecast and performance measured over a corresponding 21-day occupancy forecast.

We produced probability integral transform (PIT) plots to evaluate the calibration of the forecast (Supp. Performance, Figure 1). Calibration refers to the concordance between the distribution of our forecasts and the eventual distribution of observations [18]; for example, in a well calibrated forecast, each decile across the distribution of all forecast predictions should contain approximately 10% of the eventual observations. Where overlapping intervals contained the eventual observation (typically due to small integer counts, e.g. in smaller population size jurisdictions), we have counted each overlapping interval as containing the observation, with these down-weighted such that any given observation only contributed a total count of one.

Version control repositories are available on GitHub for the simulation and inference steps (http://github.com/ruarai/curvemush), the forecasting pipeline (http://github. com/ruarai/clinical_forecasts), and performance evaluation and manuscript figure plotting code (http://github.com/ruarai/clinical_forecasting_paper). Analysis was performed in the R statistical computing environment (version 4.3.2) [40]. The forecasting pipeline was implemented using the targets package [25], with tidyverse packages used for data manipulation [50], pracma for numerical solutions of the maximum-likelihood estimates [1], and Rcpp for interfacing with the stochastic simulation C++ code.

Forecasting performance was evaluated using the fabletools, tsibble and distributional packages [33, 49, 34].

## Data and code availability

All code is available archived at OSF (http://osf.io/5e6ma/, DOI: 10.17605/OSF.IO/5E6MA). Changes to the model which occurred throughout the study period (which was limited to jurisdiction-specific modifications to 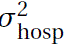 and 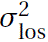 and a correction for New South Wales case data not including cases detected via rapid antigen test) are described in the supplementary materials (Supp. Methods S5).

Limited data for reproducing the figures presented in this manuscript are also available at OSF (http://osf.io/5e6ma/, DOI: 10.17605/OSF.IO/5E6MA); this includes all model output forecast trajectories, reported occupancy counts retrieved from covid19data.com.au [32], case forecast performance metrics, and Approximate Bayesian Computation diagnostic plots as produced in the course of producing occupancy forecasts. The complete line listed case dataset is not publicly available; for access to the raw data, a request must be submitted to the Australian Government Department of Health and Aged Care which will be assessed by a data committee independent of authorship group.

## Supporting information

Supplementary Forecast Performance

## Data Availability

All code is available archived at OSF (http://osf.io/5e6ma/, DOI: 10.17605/OSF.IO/5E6MA). Changes to the model which occurred throughout the study period (which was limited to jurisdiction-specific modifications to sigma_hosp and sigma_los and a correction for New South Wales case data not including cases detected via rapid antigen test) are described in the supplementary materials (Supp.\ Methods S5).
Limited data for reproducing the figures presented in this manuscript are also available at OSF (http://osf.io/5e6ma/, DOI: 10.17605/OSF.IO/5E6MA); this includes all model output forecast trajectories, reported occupancy counts retrieved from covid19data.com.au, case forecast performance metrics, and Approximate Bayesian Computation diagnostic plots as produced in the course of producing occupancy forecasts. The complete line listed case dataset is not publicly available; for access to the raw data, a request must be submitted to the Australian Government Department of Health and Aged Care which will be assessed by a data committee independent of authorship group.

## Acknowledgements

Our forecasts used surveillance data reported through the Communicable Diseases Network Australia by the interim Australian CDC (previously the Office of Health Protection), Department of Health and Aged Care on behalf of the Communicable Diseases Network Australia (CDNA) as part of the nationally coordinated response to COVID-19. We additionally used surveillance data provided by the New South Wales Department of Health. We thank public health staff in state and territory health departments, the Australian Government Department of Health and Aged Care, and in state and territory public health laboratories. We thank members of CDNA for their feedback and perspectives on the results of the analyses. This work was directly funded by the Australian Government Department of Health and Aged Care. Additional support was provided by the National Health and Medical Research Council of Australia through its Investigator Grant Schemes (FMS Emerging Leader Fellowship, 2021/GNT2010051).

## Ethics

The study was undertaken as urgent public health action to support Australia’s COVID-19 pandemic response. The study used data from the Australian National Notifiable Disease Surveillance System (NNDSS) provided to the Australian Government Department of Health and Aged Care under the National Health Security Agreement for the purposes of national communicable disease surveillance. Non-identifiable data from the NNDSS were supplied to the investigator team for the purposes of provision of epidemiological advice to government; data were securely managed to ensure patient privacy and to ensure the study’s compliance with the National Health and Medical Research Council’s Ethical Considerations in Quality Assurance and Evaluation Activities. Contractual obligations established strict data protection protocols agreed between the University of Melbourne and sub-contractors and the Australian Government Department of Health and Aged Care, with oversight and approval for use in supporting Australia’s pandemic response and for publication provided by the data custodians represented by the Communicable Diseases Network of Australia. The use of these data for these purposes, including publication, was agreed by the Department of Health with the Communicable Diseases Network of Australia. Ethical approval for this study was also provided by The University of Melbourne’s Human Research Ethics Committee (2024-26949-50575-3).

The study used routinely collected patient administration data from the New South Wales (NSW) Patient Flow Portal (PFP). De-identified PFP data were securely managed to ensure patient privacy and to ensure the study’s compliance with the National Health and Medical Research Council’s Ethical Considerations in Quality Assurance and Evaluation Activities. These data were provided for use in this study to support public health response under the governance of Health Protection NSW. The NSW Public Health Act (2010) allows for such release of data to identify and monitor risk factors for diseases and conditions that have a substantial adverse impact on the population and to improve service delivery. Following review, the NSW Ministry of Health determined that this study met that threshold and therefore provided approval for the study to proceed. Approval for publication was provided by the NSW Ministry of Health.

## Results

### Visual performance assessments

We examined the qualitative performance of our ward and ICU forecasts through visual assessment using the state of New South Wales as a case study (Figure 3). The early growth phase of BA.2 in late March and early April 2022 (forecasts 1–3) was well predicted across both ward and ICU counts. We see that the peak in ward occupancy induced by the BA.2 lineage in April was well predicted, with forecasts produced 2–3 weeks prior to the peak (forecasts 4, 5) capturing the peak in their central densities. Forecasts of the declining phase following the BA.2 peak had varied performance: the first forecast in mid-April (forecast 6) underpredicted ward occupancy, though not ICU occupancy; this was followed by two forecasts (forecasts 7, 8) with good predictive performance of ward occupancy (though forecast 8 predicted ICU occupancy with too little uncertainty); and the subsequent forecast produced in early May (forecast 9) incorrectly predicted that ward and ICU occupancy counts would increase again rather than continue to very slowly decline.

New South Wales forecasts produced during the inter-epidemic period between the BA.2 and BA.4/5 waves in late May and early June underpredicted ward occupancy and marginally underpredicted ICU occupancy (forecasts 11–14). The early growth phase of the BA.4/5 wave was not captured in our predictions until late June (forecast 15), almost a month after occupancy had begun to stabilise and then slowly increase. Similar to the BA.2 peak, early forecasts showed very good performance in terms of predicting the magnitude of the BA.4/5 peak in ward occupancy in mid-July (forecasts 17–18). However, these forecasts failed to predict the timing of the peak, instead predicting that ward occupancy would continue to increase into August. Our forecasts only correctly predicted reductions in the occupancy counts once counts had already begun to stabilise in late July (forecasts 19–21), though these still marginally over-predicted ward occupancy counts.

Further plots for the visual assessment of forecast performance for all other jurisdictions are available in the supplementary materials (Supp. Performance Figures 5–20).

### Quantitative performance

Measured forecast performance varied over the duration of the study period and across Australia’s eight states and territories (Figure 4). Measuring performance aggregated by forecast horizon (Figure 4A) shows the performance of the ward occupancy forecasts generally degraded the further into the future predictions were made. Ward occupancy performance for the Northern Territory was particularly unstable across all days of the horizon (Figure 4A). The drop in forecast performance as forecast horizon increased was less visible for the ICU forecasts (Figure 4C), likely reflecting the reduced scale of variation in the ICU time series, where the effect of changes in epidemic activity were less visible.

Median ward occupancy forecast performance averaged across all horizons was best in New South Wales (Figure 4B), possibly reflecting the use of hospital length of stay estimates derived from New South Wales data, which may not be generalisable to other jurisdictions. ICU occupancy forecast performance was best in Victoria, followed by New South Wales. The states and territories with smaller populations (Tasmania, the Australian Capital Territory and the Northern Territory) tended to have worse performance for both ward and ICU occupancy forecasts, possibly due to a greater impact of individual-level variation in length of stay where admission counts were low (Supp. Performance, Figures 5, 6, 9, 10, 15, 16). Although South Australia had a (marginally) worse median ward occupancy forecast CRPS than New South Wales (Figure 4B), examining performance across the 15–21 day horizon (Figure 4A), the CRPS for South Australia exhibited a greater consistency in performance.

Examining changes in performance of the ward occupancy forecast over the duration of the study period (Figure 5), we note associations between forecast performance and the epidemiological context, with ward occupancy forecasts often biased downwards during pre-epidemic peak phases, and biased upwards during the post-epidemic peak phases. Results for ICU occupancy forecast performance over time (Supp. Performance, Figure 2) show similar trends, though here variation in length of stay at the individual-scale likely has a greater influence on performance, given the low (<50) counts for occupancy across most jurisdictions over the study period.

We examined how the performance of the ensemble case forecast used as input to our model affected the performance of our ward and ICU forecasts. Averaged across the horizon of each of the forecasts, the mean ward forecast CRPS tended to be lower than that of the corresponding case forecast (Figure 6). This is expected given that the case forecast is a forecast of incidence, whereas our forecasts are of occupancy (i.e. prevalence), and as such exhibit greater autocorrelation and hence predictability. Comparing the ICU forecast performance to that of the case forecast (Supp. Performance, Figure 3) yields broadly similar results, although ICU performance in the Australian Capital Territory notably underperforms in comparison to the case forecasts. Bias in the case forecasts tended to be reflected in the ward occupancy forecasts (Supp. Performance, Figure 4), although this effect is less clear in jurisdictions with smaller populations such as the Northern Territory (which has a population of approximately 250,000, compared to 8.1 million for New South Wales or 6.5 million for Victoria).

In our probability integral transform plots (Supp. Performance, Figure 1), we observe that forecast calibration varies from good to poor between states and across the ward and ICU forecasts. Calibration was best for the ward forecasts in South Australia and best for the ICU forecasts in New South Wales. A few forecasts were overconfident, with Northern Territory, Queensland and Tasmanian ward forecasts and Queensland ICU forecasts having a substantial proportion of observations occurring in the bottom-or top-most intervals. A similar pattern can be observed for the New South Wales ward occupancy forecasts, with a large proportion of observations falling in the top-most interval; this was likely a consequence of a string of underpredicting forecasts from late May through to early July (Figure 3, forecasts 11–14). The ICU forecasts for South Australia and Victoria had excessive levels of uncertainty, with few observations falling in the outer intervals.

## Discussion

We have presented a clinical forecasting model for forecasting the number of patients with COVID-19 in ward and ICU beds. The model simulates the progression of patients through a compartmental model of hospital pathways, with simulations informed by near-real-time epidemiological data and fit to reported bed occupancy counts using Approximate Bayesian Computation. We have evaluated the performance of our forecasting methodology as it was applied and reported to public health decision-makers in the Australian context between March and September 2022 (although forecast outputs were produced between December 2021 and March 2022, we do not consider them in this study as the model received intensive development throughout that period). Our use of an independently produced case forecast as input to the clinical model has allowed us to take advantage of diverse case forecasting methodologies, and we have shown how the performance of our clinical forecasts can be evaluated in terms of the input case forecast performance.

Our results show that forecasting performance was variable over the study period and dependent upon the epidemiological context. The 15–21 day performance of the ward forecasts was poorest across most jurisdictions during the transition from Omicron BA.2 dominance to Omicron BA.4/5 dominance between May and July 2022 (Figure 5). This reduced performance can be observed in New South Wales from late May until early June (Figure 3, forecasts 11–14); by late June (forecast 15) a BA.4/5 transmission advantage was included in the mechanistic case forecasting models [42], increasing median predicted occupancy counts but also the uncertainty across these predictions. Forecasting during such variant transition events has previously been noted to be challenging [44].

Accurate prediction near epidemic peaks has previously been recognised to be a particularly difficult problem, both in the context of case incidence forecasts [5, 41, 4] and hospital burden forecasts [44, 28]. In our results, forecasting performance around epidemic peaks varied. Prior to peaks (in the epidemic growth phase) our forecasts generally performed well, although they tended to be biased downwards (Figure 5). Examining forecasts with start dates in the weeks prior to epidemic peaks (Supp. Performance, Figures 5–20), we see that occupancy count at the peak was generally well captured by forecasts produced one or two weeks prior to the point of peak occupancy. Forecasts which were produced three weeks prior to the peak performed worse, with most predicting that occupancy would continue to grow beyond what eventuated to be the peak. However, at this three-week horizon, the forecasts typically had wide credible intervals which appropriately conveyed the uncertainty of our predictions.

Our use of an independently produced forecast of case incidence as input distinguishes our work from previously published methods for forecasting COVID-19 clinical burden, which often simultaneously model both infection dynamics and the subsequent pathways taken by infected patients through the hospital system [51, 35, 24, 16] or produce statistical predictions of occupancy without incorporation of causal mechanisms [37, 16]. The decoupling of our clinical progression model from the case forecasting models allows for greater ‘separation of concerns’ since the development of each model can occur independently [26]. Further, Figure 6 demonstrates that the quality of our occupancy forecasts depends upon the performance of the input case incidence forecasts (a similar result has been previously reported for a model of hospital admissions [29]), implying that our use of an ensemble case forecast as input has been advantageous for the performance of our occupancy forecasts, given ensembles have repeatedly been shown to improve case forecasting performance [38, 12, 36, 29, 44].

Our clinical forecasting model is designed to receive outputs from forecasts of case incidence as a (large) sample of trajectories. However, it has been more common for forecast outputs to be summarised using prediction intervals which quantify the probability of outcomes falling within certain ranges. Examples of this have included the collaborative ensemble forecasts reported by the US and European COVID-19 forecast hubs [11, 44]. These prediction intervals are incompatible with our methodology as they obscure the underlying autocorrelation in the case incidence time series — if we were to sample from such intervals across each day of the forecast, uncertainty in the cumulative case count would be underestimated. We recommend that collaborative ensemble forecasts of infectious disease report outputs as trajectories where possible, so as to enable the appropriate propagation of uncertainty in further applications (such as that presented here).

Infectious disease forecasting models often exhibit reduced performance when predicting in low count contexts [37, 35]. In our work, we produced forecasts across low counts of both ward and ICU occupancy, typically during inter-epidemic periods and in jurisdictions with smaller population sizes. The performance of our forecasts as measured through CRPS was worse in these contexts (Figure 4). However, this is in large part due to the CRPS being calculated over log-transformed counts — effectively making it a measure of relative error. This would be expected to penalise forecasts produced in low count contexts, where small absolute changes can produce large relative differences [3]. We also note that the performance of our occupancy forecasts across these low count contexts may be of lesser importance to public health decision-makers given they are typically (by definition) distant from capacity constraints.

Since the clinical forecasting model is informed by near-real-time estimates of key quantities such as probability of hospitalisation and length of stay, reasonable forecast performance could be expected in the absence of the approximate Bayesian fitting step. While this occasionally proved to be true in application (e.g., see Supp. Methods Figure 1, where the model output without fitting captures the reported ward occupancy counts for the Northern Territory), a few factors may have prevented this from being generally the case: firstly, we used patient length of stay distributions which were fit to data from the state of New South Wales and these distributions may not reflect the clinical practice or realised severity in other jurisdictions; secondly, the compartmental model we used may miss some components of hospital occupancy dynamics, such as outbreaks of COVID-19 within hospitals; thirdly, we assumed that the population which was reported as hospitalised in the case data was the same population as that reported in the hospital occupancy figures, which was not always the case due to differing upstream datasets (e.g. Victoria collected occupancy counts as a separate census of patients [47]); finally, our near-real-time estimates of ward and ICU admission probability were not adjusted for possible right-truncation due to reporting lags as the date of data entry was not available within the case dataset we had access to.

The measure of hospital burden we chose to forecast — hospital occupancy — has an advantage over incidence measures such as daily hospital admissions since it directly relates to the capacity of the healthcare system. However, it has a few disadvantages of note. Because hospital occupancy is a prevalence measure, it is inherently slower to respond to changes in the epidemic situation than hospital admissions and is therefore less useful as an indicator of changes in epidemic activity. It may also be more difficult to measure at the hospital level, given it requires either accurate accounting of admissions and discharges or recording of individual patient stays. Ideally, both admissions and occupancy would be monitored and reported; in such a context our model could be easily extended to fit to and report admission counts, given admission counts are already recorded within our simulations.

Throughout the period for which COVID-19 bed occupancy counts were collected and reported in Australia, no nationally consistent standard specified which COVID-19 cases should be included in the counts. As a result, distinct definitions were created and applied across jurisdictions. For example, during our study period, the state of New South Wales counted any patient in hospital who had been diagnosed with COVID-19 either during their hospital stay or within the 14 days prior to their admission to hospital [21]. This broad definition had the beneficial effect of reducing false-negatives in the counting process but resulted in the inclusion of a large number of individuals who had since recovered from infection and/or whose stay was unrelated to the disease (with this effect then being captured in the estimates of length of stay used in our study). This was in contrast to Victoria, where COVID-19 cases were counted only until a negative test result was received [47], reducing false positive inclusions but underestimating the total hospital burden of the disease, given COVID-19 cases may still require hospital care or be isolated for infection control reasons even when they no longer test positive. Although these differences would not be expected to substantially affect the forecast performance given our fitting methodology, the development of standard definitions that could be applied in future epidemics would allow for direct comparison of counts between jurisdictions and simplify modelling efforts.

The modelling framework we have described here is flexible and not inherently tied to COVID-19 hospital occupancy as the forecasting target. In general terms, our method stochastically simulates the convolution of a time series of case incidence into time series of subsequent outcomes. As such, the methodology could be applied to measures of other infectious diseases. Further, the framework could be used to model other outcomes of infectious disease, such as workforce absenteeism or long-term sequelae.

We have presented a robust approach for forecasting COVID-19 hospital ward and ICU bed occupancy and have examined the performance of this methodology as applied in the Australian context between March and September 2022. Our approach takes as input an independently produced forecast of case incidence and, in combination with near-realtime estimates of epidemiological parameters, simulates the progression of patients from case onset through to hospital ward and ICU care. Our use of independently produced forecasts of case incidence has allowed us to both develop our model independently of the input case forecasting models and take advantage of the performance benefits provided by ensemble case forecasts. Our computationally efficient inference method allowed us to generate forecasts for multiple Australian jurisdictions in near-real-time, enabling the rapid provision of evidence to public health decision-makers.

## References

[1] Hans W. Borchers. pracma: Practical Numerical Math Functions, 2023. R package version 2.4.4.

[2] Nikos I Bosse, Sam Abbott, Anne Cori, Edwin van Leeuwen, Johannes Bracher, and Sebastian Funk. Scoring epidemiological forecasts on transformed scales. PLoS Comput. Biol., 19(8):e1011393, August 2023.

[3] Nikos I Bosse, Sam Abbott, Anne Cori, Edwin van Leeuwen, Johannes Bracher, and Sebastian Funk. Transformation of forecasts for evaluating predictive performance in an epidemiological context. bioRxiv, January 2023.

[4] J Bracher, D Wolffram, J Deuschel, K Görgen, J L Ketterer, A Ullrich, S Abbott, M V Barbarossa, D Bertsimas, S Bhatia, M Bodych, N I Bosse, J P Burgard, L Castro, G Fairchild, J Fuhrmann, S Funk, K Gogolewski, Q Gu, S Heyder, T Hotz, Y Kheifetz, H Kirsten, T Krueger, E Krymova, M L Li, J H Meinke, I J Michaud, K Niedzielewski, T Ożański, F Rakowski, M Scholz, S Soni, A Srivastava, J Zieliński, D Zou, T Gneiting, M Schienle, and List of Contributors by Team. A pre-registered short-term forecasting study of COVID-19 in Germany and Poland during the second wave. Nat. Commun., 12(1):5173, August 2021.

[5] Mario Castro, Saúl Ares, José A Cuesta, and Susanna Manrubia. The turning point and end of an expanding epidemic cannot be precisely forecast. Proc. Natl. Acad. Sci. U. S. A., 117(42):26190–26196, October 2020.

[6] Allen C Cheng, Dominic E Dwyer, Mark Holmes, Louis Irving, Graham Simpson, Sanjaya Senenayake, Tony Korman, N Deborah Friedman, Louise Cooley, Peter Wark, Anna Holwell, Simon Bowler, John Upham, Daniel M Fatovich, Grant Waterer, Christopher C Blyth, Nigel Crawford, Jim Buttery, Helen S Marshall, Julia E Clark, Joshua Francis, Kristine Macartney, Tom Kotsimbos, and Paul Kelly. Influenza epidemiology in patients admitted to sentinel Australian hospitals in 2019: the Influenza Complications Alert Network (FluCAN). Commun. Dis. Intell., 46, April 2022.

[7] Communicable Diseases Network Australia. Australian National Disease Surveillance Plan for COVID-19. Technical report, Australian Government Department of Health, 2022.

[8] Eamon Conway, Camelia R Walker, Christopher Baker, Michael J Lydeamore, Gerard E Ryan, Trish Campbell, Joel C Miller, Nic Rebuli, Max Yeung, Greg Kabashima, Nicholas Geard, James Wood, James M McCaw, Jodie McVernon, Nick Golding, David J Price, and Freya M Shearer. COVID-19 vaccine coverage targets to inform reopening plans in a low incidence setting. Proceedings of the Royal Society B, August 2023.

[9] COVID-19 Epidemiology and Surveillance Team. COVID-19 Australia: Epidemiology Report 67 Reporting period ending 23 October 2022. Commun. Dis. Intell., 46, November 2022.

[10] COVID-19 National Incident Room Surveillance Team. COVID-19 Australia: Epidemiology Report 62 Reporting period ending 5 June 2022. Commun. Dis. Intell., 46, July 2022.

[11] Estee Y Cramer, Yuxin Huang, Yijin Wang, Evan L Ray, Matthew Cornell, Johannes Bracher, Andrea Brennen, Alvaro J Castro Rivadeneira, Aaron Gerding, Katie House, Dasuni Jayawardena, Abdul Hannan Kanji, Ayush Khandelwal, Khoa Le, Vidhi Mody, Vrushti Mody, Jarad Niemi, Ariane Stark, Apurv Shah, Nutcha Wattanchit, Martha W Zorn, Nicholas G Reich, and US COVID-19 Forecast Hub Consortium. The United States COVID-19 Forecast Hub dataset. Sci. Data, 9(1):462, August 2022.

[12] Estee Y Cramer, Evan L Ray, Velma K Lopez, Johannes Bracher, Andrea Brennen, Alvaro J Castro Rivadeneira, Aaron Gerding, Tilmann Gneiting, Katie H House, Yuxin Huang, Dasuni Jayawardena, Abdul H Kanji, Ayush Khandelwal, Khoa Le, Anja Mühlemann, Jarad Niemi, Apurv Shah, Ariane Stark, Yijin Wang, Nutcha Wattanachit, Martha W Zorn, Youyang Gu, Sansiddh Jain, Nayana Bannur, Ayush Deva, Mihir Kulkarni, Srujana Merugu, Alpan Raval, Siddhant Shingi, Avtansh Tiwari, Jerome White, Neil F Abernethy, Spencer Woody, Maytal Dahan, Spencer Fox, Kelly Gaither, Michael Lachmann, Lauren Ancel Meyers, James G Scott, Mauricio Tec, Ajitesh Srivastava, Glover E George, Jeffrey C Cegan, Ian D Dettwiller, William P England, Matthew W Farthing, Robert H Hunter, Brandon Lafferty, Igor Linkov, Michael L Mayo, Matthew D Parno, Michael A Rowland, Benjamin D Trump, Yanli Zhang-James, Samuel Chen, Stephen V Faraone, Jonathan Hess, Christopher P Morley, Asif Salekin, Dongliang Wang, Sabrina M Corsetti, Thomas M Baer, Marisa C Eisenberg, Karl Falb, Yitao Huang, Emily T Martin, Ella McCauley, Robert L Myers, Tom Schwarz, Daniel Sheldon, Graham Casey Gibson, Rose Yu, Liyao Gao, Yian Ma, Dongxia Wu, Xifeng Yan, Xiaoyong Jin, Yu-Xiang Wang, Yangquan Chen, Lihong Guo, Yanting Zhao, Quanquan Gu, Jinghui Chen, Lingxiao Wang, Pan Xu, Weitong Zhang, Difan Zou, Hannah Biegel, Joceline Lega, Steve McConnell, V P Nagraj, Stephanie L Guertin, Christopher Hulme-Lowe, Stephen D Turner, Yunfeng Shi, Xuegang Ban, Robert Walraven, Qi-Jun Hong, Stanley Kong, Axel van de Walle, James A Turtle, Michal Ben-Nun, Steven Riley, Pete Riley, Ugur Koyluoglu, David DesRoches, Pedro Forli, Bruce Hamory, Christina Kyriakides, Helen Leis, John Milliken, Michael Moloney, James Morgan, Ninad Nirgudkar, Gokce Ozcan, Noah Piwonka, Matt Ravi, Chris Schrader, Elizabeth Shakhnovich, Daniel Siegel, Ryan Spatz, Chris Stiefeling, Barrie Wilkinson, Alexander Wong, Sean Cavany, Guido España, Sean Moore, Rachel Oidtman, Alex Perkins, David Kraus, Andrea Kraus, Zhifeng Gao, Jiang Bian, Wei Cao, Juan Lavista Ferres, Chaozhuo Li, Tie-Yan Liu, Xing Xie, Shun Zhang, Shun Zheng, Alessandro Vespignani, Matteo Chinazzi, Jessica T Davis, Kunpeng Mu, Ana Pastore Y Piontti, Xinyue Xiong, Andrew Zheng, Jackie Baek, Vivek Farias, Andreea Georgescu, Retsef Levi, Deeksha Sinha, Joshua Wilde, Georgia Perakis, Mohammed Amine Bennouna, David Nze-Ndong, Divya Singhvi, Ioannis Spantidakis, Leann Thayaparan, Asterios Tsiourvas, Arnab Sarker, Ali Jadbabaie, Devavrat Shah, Nicolas Della Penna, Leo A Celi, Saketh Sundar, Russ Wolfinger, Dave Osthus, Lauren Castro, Geoffrey Fairchild, Isaac Michaud, Dean Karlen, Matt Kinsey, Luke C Mullany, Kaitlin Rainwater-Lovett, Lauren Shin, Katharine Tallaksen, Shelby Wilson, Elizabeth C Lee, Juan Dent, Kyra H Grantz, Alison L Hill, Joshua Kaminsky, Kathryn Kaminsky, Lindsay T Keegan, Stephen A Lauer, Joseph C Lemaitre, Justin Lessler, Hannah R Meredith, Javier Perez-Saez, Sam Shah, Claire P Smith, Shaun A Truelove, Josh Wills, Maximilian Marshall, Lauren Gardner, Kristen Nixon, John C Burant, Lily Wang, Lei Gao, Zhiling Gu, Myungjin Kim, Xinyi Li, Guannan Wang, Yueying Wang, Shan Yu, Robert C Reiner, Ryan Barber, Emmanuela Gakidou, Simon I Hay, Steve Lim, Chris Murray, David Pigott, Heidi L Gurung, Prasith Baccam, Steven A Stage, Bradley T Suchoski, B Aditya Prakash, Bijaya Adhikari, Jiaming Cui, Alexander Rodríguez, Anika Tabassum, Jiajia Xie, Pinar Keskinocak, John Asplund, Arden Baxter, Buse Eylul Oruc, Nicoleta Serban, Sercan O Arik, Mike Dusenberry, Arkady Epshteyn, Elli Kanal, Long T Le, Chun-Liang Li, Tomas Pfister, Dario Sava, Rajarishi Sinha, Thomas Tsai, Nate Yoder, Jinsung Yoon, Leyou Zhang, Sam Abbott, Nikos I Bosse, Sebastian Funk, Joel Hellewell, Sophie R Meakin, Katharine Sherratt, Mingyuan Zhou, Rahi Kalantari, Teresa K Yamana, Sen Pei, Jeffrey Shaman, Michael L Li, Dimitris Bertsimas, Omar Skali Lami, Saksham Soni, Hamza Tazi Bouardi, Turgay Ayer, Madeline Adee, Jagpreet Chhatwal, Ozden O Dalgic, Mary A Ladd, Benjamin P Linas, Peter Mueller, Jade Xiao, Yuanjia Wang, Qinxia Wang, Shanghong Xie, Donglin Zeng, Alden Green, Jacob Bien, Logan Brooks, Addison J Hu, Maria Jahja, Daniel McDonald, Balasubramanian Narasimhan, Collin Politsch, Samyak Rajanala, Aaron Rumack, Noah Simon, Ryan J Tibshirani, Rob Tibshirani, Valerie Ventura, Larry Wasserman, Eamon B O’Dea, John M Drake, Robert Pagano, Quoc T Tran, Lam Si Tung Ho, Huong Huynh, Jo W Walker, Rachel B Slayton, Michael A Johansson, Matthew Biggerstaff, and Nicholas G Reich. Evaluation of individual and ensemble probabilistic forecasts of COVID-19 mortality in the United States. Proc. Natl. Acad. Sci. U. S. A., 119(15):e2113561119, April 2022.

[13] Christopher R Dale, Rachael W Starcher, Shu Ching Chang, Ari Robicsek, Guilford Parsons, Jason D Goldman, Andre Vovan, David Hotchkin, and Tyler J Gluckman. Surge effects and survival to hospital discharge in critical care patients with COVID-19 during the early pandemic: a cohort study. Crit. Care, 25(1):70, February 2021.

[14] Department of Health and Aged Care. Australia’s COVID-19 Vaccine Rollout. Technical report, August 2022.

[15] Kevin J Fong, Charlotte Summers, and Tim M Cook. NHS hospital capacity during covid-19: overstretched staff, space, systems, and stuff. BMJ, 385:e075613, April 2024.

[16] S Funk, S Abbott, B D Atkins, M Baguelin, J K Baillie, P Birrell, J Blake, N I Bosse, J Burton, J Carruthers, N G Davies, D De Angelis, L Dyson, W J Edmunds, R M Eggo, N M Ferguson, K Gaythorpe, E Gorsich, G Guyver-Fletcher, J Hellewell, E M Hill, A Holmes, T A House, C Jewell, M Jit, T Jombart, I Joshi, M J Keeling, E Kendall, E S Knock, A J Kucharski, K A Lythgoe, S R Meakin, J D Munday, P J M Openshaw, C E Overton, F Pagani, J Pearson, P N Perez-Guzman, L Pellis, F Scarabel, M G Semple, K Sherratt, M Tang, M J Tildesley, E Van Leeuwen, L K Whittles, CMMID COVID-19 Working Group, Imperial College COVID-19 Response Team, and ISARIC4C Investigators. Short-term forecasts to inform the response to the Covid-19 epidemic in the UK. bioRxiv, November 2020.

[17] Sebastian Funk, Anton Camacho, Adam J Kucharski, Rachel Lowe, Rosalind M Eggo, and W John Edmunds. Assessing the performance of real-time epidemic forecasts: A case study of Ebola in the Western Area region of Sierra Leone, 2014-15. PLoS Comput. Biol., 15(2):e1006785, February 2019.

[18] Tilmann Gneiting, Fadoua Balabdaoui, and Adrian E Raftery. Probabilistic forecasts, calibration and sharpness. J. R. Stat. Soc. Series B Stat. Methodol., 69(2):243–268, April 2007.

[19] Nick Golding, Tianxiao Hao, Gerry Ryan, Adeshina Adekunle, Peter Dawson, Mingmei Teo, Robert Moss, David J Price, Freya M Shearer, Ruarai Tobin, McCaw, James M., Dyland Morris, Ross, Joshua V., Tobin South, Rob Hyndman, Michael Lydeamore, Mitchell O‘Hara-Wild, and James Wood. Situational assessment of COVID-19 in Australia — Technical Report 22 May 2022. Technical report, August 2022.

[20] Nick Golding, David J Price, Gerard Ryan, Jodie McVernon, James M McCaw, and Freya M Shearer. A modelling approach to estimate the transmissibility of SARS-CoV-2 during periods of high, low, and zero case incidence. eLife, 12, January 2023.

[21] Health Protection New South Wales. New South Wales COVID-19 weekly data overview, epidemiological week 9. Technical report, March 2022.

[22] Rob Hyndman and George Athanasopoulos. Forecasting: Principles and Practice (3rd ed). OTexts, 2021.

[23] Sameer S Kadri, Junfeng Sun, Alexander Lawandi, Jeffrey R Strich, Lindsay M Busch, Michael Keller, Ahmed Babiker, Christina Yek, Seidu Malik, Janell Krack, John P Dekker, Alicen B Spaulding, Emily Ricotta, John H Powers, 3rd, Chanu Rhee, Michael Klompas, Janhavi Athale, Tegan K Boehmer, Adi V Gundlapalli, William Bentley, S Deblina Datta, Robert L Danner, Cumhur Y Demirkale, and Sarah Warner. Association between caseload surge and COVID-19 survival in 558 U.S. hospitals, March to August 2020. Ann. Intern. Med., 174(9):1240–1251, September 2021.

[24] D Klinkenberg, J Backer, N de Keizer, S McDonald, and J Wallinga. Projecting COVID-19 intensive care admissions in the Netherlands for policy advice: February 2020 to January 2021. Technical report, National Institute for Public Health and the Environment, The Netherlands, March 2023.

[25] William Michael Landau. The targets r package: a dynamic make-like function-oriented pipeline toolkit for reproducibility and high-performance computing. Journal of Open Source Software, 6(57):2959, 2021.

[26] Philip A Laplante. What every engineer should know about software engineering. What Every Engineer Should Know. CRC Press, Boca Raton, FL, April 2007.

[27] Dorothy Machalek, Eithandee Aung, John Kaldor, Noni Winkler, Heather Gidding, Kristine Macartney, Rena Hirani, Perfecto Diaz, Mark Sombillo, Sue Ismay, Haydon Maas, Kirsty Pevitt, Katrina Smith, Iain Gosbell, David Irving, Suellen Nicholson, Theo Karapanagiotidis, Rianne Brizuela, Natalie Cain, Han Huang, Karla Hernandez, Deborah Williamson, John Carlin, and Matthew O’sullivan. Seroprevalence of SARS-CoV-2-specific antibodies among Australian blood donors, February-March 2022. Technical report, 2022.

[28] Harrison Manley, Thomas Bayley, Gabriel Danelian, Lucy Burton, Thomas Finnie, André Charlett, Nicholas A Watkins, Paul Birrel, Daniela De Angelis, Matt Keeling, Sebastian Funk, Graham F Medley, Lorenzo Pellis, Marc Baguelin, Graeme J Ackland, Johanna Hutchinson, Steven Riley, and Jasmina Panovska-Griffiths. Combining models to generate consensus medium-term projections of hospital admissions, occupancy and deaths relating to COVID-19 in England. bioRxiv, November 2023.

[29] Sophie Meakin, Sam Abbott, Nikos Bosse, James Munday, Hugo Gruson, Joel Hellewell, Katharine Sherratt, CMMID COVID-19 Working Group, and Sebastian Funk. Comparative assessment of methods for short-term forecasts of COVID-19 hospital admissions in England at the local level. BMC Med., 20(1):86, February 2022.

[30] Robert Moss, David J Price, Nick Golding, Peter Dawson, Jodie McVernon, Rob J Hyndman, Freya M Shearer, and James M McCaw. Forecasting COVID-19 activity in Australia to support pandemic response: May to October 2020. Sci. Rep., 13(1):8763, May 2023.

[31] Robert Moss, James Wood, Damien Brown, Freya M Shearer, Andrew J Black, Kathryn Glass, Allen C Cheng, James M McCaw, and Jodie McVernon. Coronavirus disease model to inform transmission-reducing measures and health system preparedness, Australia. Emerg. Infect. Dis., 26(12):2844–2853, December 2020.

[32] Juliette O’Brien, David Barry, Daniel Babekuhl, Matthew Bolton, Carlos Monteiro, Liam Pearson, Hugh Kennedy, Hamish Goodwin, Joe Garcia, and Louise Lor. covid19data.com.au. https://www.covid19data.com.au/. Accessed: –.

[33] Mitchell O’Hara-Wild, Rob Hyndman, and Earo Wang. fabletools: Core Tools for Packages in the ‘fable’ Framework, 2023. R package version 0.3.4.

[34] Mitchell O’Hara-Wild, Matthew Kay, and Alex Hayes. distributional: Vectorised Probability Distributions, 2023. R package version 0.3.2.

[35] Christopher E Overton, Lorenzo Pellis, Helena B Stage, Francesca Scarabel, Joshua Burton, Christophe Fraser, Ian Hall, Thomas A House, Chris Jewell, Anel Nurtay, Filippo Pagani, and Katrina A Lythgoe. EpiBeds: Data informed modelling of the COVID-19 hospital burden in England. PLoS Comput. Biol., 18(9):e1010406, September 2022.

[36] Juliette Paireau, Alessio Andronico, Nathanaël Hozé, Maylis Layan, Pascal Crépey, Alix Roumagnac, Marc Lavielle, Pierre-Yves Boëlle, and Simon Cauchemez. An ensemble model based on early predictors to forecast COVID-19 health care demand in France. Proc. Natl. Acad. Sci. U. S. A., 119(18):e2103302119, May 2022.

[37] Mark J Panaggio, Kaitlin Rainwater-Lovett, Paul J Nicholas, Mike Fang, Hyunseung Bang, Jeffrey Freeman, Elisha Peterson, and Samuel Imbriale. Gecko: A time-series model for COVID-19 hospital admission forecasting. Epidemics, 39(100580):100580, June 2022.

[38] Pierre Pinson. Comparing ensemble approaches for short-term probabilistic COVID-19 forecasts in the U.S. International Institute of Forecasters. https://forecasters.org/blog/2020/10/28/comparing-ensemble-approaches-for-short-term-probabilistic-covid-19-forecasts-in-the-u-s/, October 2020. Accessed: 2023-7-10.

[39] David J Price, Freya M Shearer, Michael T Meehan, Emma McBryde, Robert Moss, Nick Golding, Eamon J Conway, Peter Dawson, Deborah Cromer, James Wood, Sam Abbott, Jodie McVernon, and James M McCaw. Early analysis of the Australian COVID-19 epidemic. Elife, 9, August 2020.

[40] R Core Team. R: A Language and Environment for Statistical Computing. R Foundation for Statistical Computing, Vienna, Austria, 2023.

[41] Nicholas G Reich, Ryan J Tibshirani, Evan L Ray, and Roni Rosenfeld. On the predictability of COVID-19. https://forecasters.org/blog/2021/09/28/on-the-predictability-of-covid-19/, September 2021. Accessed: 2024-2-23.

[42] Freya Shearer, Adekunle Adeshina, Peter Dawson, Nick Golding, Tianxiao Hao, Rob Hyndman, Rob Moss, Mitchell O’Hara-Wild, David Price, Gerard Ryan, Katharine Senior, Ruarai Tobin, James Wood, and James Mccaw. Series of weekly COVID-19 epidemic situational assessment reports submitted to the Australian Government Department of Health Office of Health Protection from April 2020 to December 2023. Technical report, 2024.

[43] Freya M Shearer, James M McCaw, Gerard E Ryan, Tianxiao Hao, Nicholas J Tierney, Michael J Lydeamore, Logan Wu, Kate Ward, Sally Ellis, James Wood, Jodie McVernon, and Nick Golding. Estimating the impact of test-trace-isolate-quarantine systems on SARS-CoV-2 transmission in Australia. Epidemics, 47(100764):100764, June 2024.

[44] Katharine Sherratt, Hugo Gruson, Rok Grah, Helen Johnson, Rene Niehus, Bastian Prasse, Frank Sandmann, Jannik Deuschel, Daniel Wolffram, Sam Abbott, Alexander Ullrich, Graham Gibson, Evan L Ray, Nicholas G Reich, Daniel Sheldon, Yijin Wang, Nutcha Wattanachit, Lijing Wang, Jan Trnka, Guillaume Obozinski, Tao Sun, Dorina Thanou, Loic Pottier, Ekaterina Krymova, Jan H Meinke, Maria Vittoria Barbarossa, Neele Leithauser, Jan Mohring, Johanna Schneider, Jaroslaw Wlazlo, Jan Fuhrmann, Berit Lange, Isti Rodiah, Prasith Baccam, Heidi Gurung, Steven Stage, Bradley Suchoski, Jozef Budzinski, Robert Walraven, Inmaculada Villanueva, Vit Tucek, Martin Smid, Milan Zajicek, Cesar Perez Alvarez, Borja Reina, Nikos I Bosse, Sophie R Meakin, Lauren Castro, Geoffrey Fairchild, Isaac Michaud, Dave Osthus, Pierfrancesco Alaimo Di Loro, Antonello Maruotti, Veronika Eclerova, Andrea Kraus, David Kraus, Lenka Pribylova, Bertsimas Dimitris, Michael Lingzhi Li, Soni Saksham, Jonas Dehning, Sebastian Mohr, Viola Priesemann, Grzegorz Redlarski, Benjamin Bejar, Giovanni Ardenghi, Nicola Parolini, Giovanni Ziarelli, Wolfgang Bock, Stefan Heyder, Thomas Hotz, David E Singh, Miguel Guzman-Merino, Jose L Aznarte, David Morina, Sergio Alonso, Enric Alvarez, Daniel Lopez, Clara Prats, Jan Pablo Burgard, Arne Rodloff, Tom Zimmermann, Alexander Kuhlmann, Janez Zibert, Fulvia Pennoni, Fabio Divino, Marti Catala, Gianfranco Lovison, Paolo Giudici, Barbara Tarantino, Francesco Bartolucci, Giovanna Jona Lasinio, Marco Mingione, Alessio Farcomeni, Ajitesh Srivastava, Pablo Montero-Manso, Aniruddha Adiga, Benjamin Hurt, Bryan Lewis, Madhav Marathe, Przemyslaw Porebski, Srinivasan Venkatramanan, Rafal P Bartczuk, Filip Dreger, Anna Gambin, Krzysztof Gogolewski, Magdalena Gruziel-Slomka, Bartosz Krupa, Antoni Moszyński, Karol Niedzielewski, Jedrzej Nowosielski, Maciej Radwan, Franciszek Rakowski, Marcin Semeniuk, Ewa Szczurek, Jakub Zielinski, Jan Kisielewski, Barbara Pabjan, Kirsten Holger, Yuri Kheifetz, Markus Scholz, Biecek Przemyslaw, Marcin Bodych, Maciej Filinski, Radoslaw Idzikowski, Tyll Krueger, Tomasz Ozanski, Johannes Bracher, and Sebastian Funk. Predictive performance of multi-model ensemble forecasts of COVID-19 across European nations. Elife, 12, April 2023.

[45] Mikael Sunnåker, Alberto Giovanni Busetto, Elina Numminen, Jukka Corander, Matthieu Foll, and Christophe Dessimoz. Approximate Bayesian computation. PLoS Comput. Biol., 9(1):e1002803, January 2013.

[46] Ruarai J Tobin, James G Wood, Duleepa Jayasundara, Grant Sara, Camelia R Walker, Genevieve E Martin, James M McCaw, Freya M Shearer, and David J Price. Real-time analysis of hospital length of stay in a mixed SARS-CoV-2 Omicron and Delta epidemic in New South Wales, Australia. BMC Infect. Dis., 23(1):28, January 2023.

[47] Victorian Department of Health and Victorian Agency for Health Information. COVID-19 Daily Capacity and Occupancy Register. Technical report, October 2021.

[48] Bruno A Walther and Joslin L Moore. The concepts of bias, precision and accuracy, and their use in testing the performance of species richness estimators, with a literature review of estimator performance. Ecography (Cop.), 28(6):815–829, December 2005.

[49] Earo Wang, Dianne Cook, and Rob J Hyndman. A new tidy data structure to support exploration and modeling of temporal data. Journal of Computational and Graphical Statistics, 29(3):466–478, 2020.

[50] Hadley Wickham, Mara Averick, Jennifer Bryan, Winston Chang, Lucy D’Agostino McGowan, Romain François, Garrett Grolemund, Alex Hayes, Lionel Henry, Jim Hester, Max Kuhn, Thomas Lin Pedersen, Evan Miller, Stephan Milton Bache, Kirill Müller, Jeroen Ooms, David Robinson, Dana Paige Seidel, Vitalie Spinu, Kohske Takahashi, Davis Vaughan, Claus Wilke, Kara Woo, and Hiroaki Yutani. Welcome to the tidyverse. Journal of Open Source Software, 4(43):1686, 2019.

[51] Cheng Zhao, Burcu Tepekule, Nicola G Criscuolo, Pedro D Wendel Garcia, Matthias P Hilty, Risc-Icu Consortium Investigators In Switzerland, Thierry Fumeaux, and Thomas Van Boeckel. icumonitoring.ch: a platform for short-term forecasting of intensive care unit occupancy during the COVID-19 epidemic in Switzerland. Swiss Med. Wkly, 150:w20277, May 2020.

