## Supplementary Methods for "A modular approach to forecasting COVID-19 hospital bed occupancy"

### Supplementary Materials — Supplementary Methods

#### Contents


1 Estimation of parameters under right-truncation 1

2 Stochastic progression model 1

3 Approximate Bayesian Computation example 3

4 Length of stay and transition probability estimates 3

5 Model changes throughout the study period 5


#### 1 Estimation of parameters under right-truncation

We calculated the probability of a case being hospitalised, $p_{\text{hosp}}(a,t)$, adjusting for right-truncation using an assumed delay from symptom onset to hospital admission. Let $F_{\text{hosp}}(t)$ be the cumulative density function for this delay duration, $d$ be the date the data was extracted, $\mathscr{A}$ be the set of individuals in the data marked as having been hospitalised, and $\mathscr{B}$ the set of individuals who have not. The likelihood of hospitalisation statuses given a probability of hospitalisation $p_{\text{hosp}}$ and dates of case symptom onset $\tau_i$ (where $\tau_i \leq d$) is then:

$$L(\mathscr{A},\mathscr{B}|p_{\text{hosp}},\tau) = \prod_{i\in\mathscr{A}} p_{\text{hosp}} F_{\text{hosp}}(d-\tau_i+1) \prod_{i\in\mathscr{B}} (1-p_{\text{hosp}}F_{\text{hosp}}(d-\tau_i+1))$$

With a corresponding score function for $p_{\text{h}}$ of:

$$S = \frac{|A|}{p_{\text{hosp}}} - \sum_{j\in\mathscr{B}} \frac{F_{\text{hosp}}(d-\tau_j+1)}{1-p_{\text{hosp}}F_{\text{hosp}}(d-\tau_j+1)},$$

such that the maximum likelihood estimate of $p_{\text{hosp}}$ is solved by setting $S$ to be zero. Note that if there is no delay (i.e. $F_{\text{hosp}}(0) = 1$), this reduces simply to the proportion of cases hospitalised $|\mathscr{A}|/(|\mathscr{A}|+|\mathscr{B}|)$.

We used the same approach for calculating the probability of a case being admitted to ICU conditional on them having been admitted to hospital, $p_{\text{ICU}}(a,t)$. In this case, $\mathscr{A}$ is instead the set of individuals who have been admitted to the ICU, and $\mathscr{B}$ is the set of individuals who have not been admitted to the ICU (but have been hospitalised). This requires the use of the delay from onset to ICU admission $F_{\text{ICU}}$; as this was not directly available, we constructed it as the delay from symptom onset to hospitalisation $F_{\text{hosp}}$ followed by the delay from hospitalisation to ICU admission $F_{\text{ICU|hosp}}$, given that a hospitalisation has occurred:

$$F_{ICU}(x) = \frac{\int_0^x F_{\text{ICU|hosp}}(x-y) f_{\text{hosp}}(y)\ dy}{F_{\text{hosp}}(x)}$$

#### 2 Stochastic progression model

For each simulation, an array $A_i$ is constructed, with a descending hierarchical structure of integer time $t \in [0, \texttt{t\_max})$, compartment $c \in [0,10]$ and 'slot' $s \in 0,1$ such that the array is indexed as $i = 11 \cdot 2 \cdot t + 2 \cdot c + s$, where $i \in [0, \texttt{t\_max} \cdot 11 \cdot 2)$. The slot value $s$ indicates where that the array either represents, for a compartment $c$ at some time $t$, the occupancy count if $s = \texttt{occupancy} = 0$ or inward transition count if $c = \texttt{transitions} = 1$. Pseudocode for simulation of the compartmental model across this structure is provided in Algorithm 1.

The hierarchical structure of the array $A_i$ allows for efficient processing of the model, reducing cache misses by minimising the across-memory distance that needs to be traversed as the simulation progresses. For each time-step, memory traversal is limited to either $t \to t+1$ (moving to the next time-step) or $t \to t+\delta_i$ (to process a transition event), with $\delta_i$ (samples from the delay distributions) expected to be relatively small to the total number of time-steps. In practice, this means the algorithm operates over a small span of the array $A_i$, reducing the rate at which uncached sections of the array need to be retrieved (i.e. reducing cache misses).

**Algorithm 1:** Stochastic compartmental progression model with arbitrary delay distributions

**Data:** Time-series of hospitalised cases $h_t$ over $D$ days
**Parameters:** $\tau$ time-steps to be simulated per day, a transition branching function sample_next_compartment and a delay sampling function sample_delays
**Result:** Occupancy and inward-transition count time-series for each compartment

```
Defining the array and array indexing function
A ⟵ ∅;
ix(t, c, s) := 11 · 2 · t + 2 · c + s;

Initialising the symptomatic inward-transition counts
for d ← 0 to D − 1 do
    A[ix(d · τ, symptomatic, transitions)] = h_d;

The maximum time-step will be number of days D by time-steps per day τ
t_max := D · τ;

The main model loop over each time-step and compartment
for t ← 0 to t_max - 1 do
    for c ← 0 to 11 − 1 do
        If not at the first time-step, continue the cumulative sum of occupancy
        if t > 0 then
            A[ix(t, c, occupancy)] = A[ix(t, c, occupancy)] + A[ix(t − 1, c, occupancy)];

        Get the number of inward transitions to handle N and add it to our occupancy count
        N = A[ix(t, c, transitions)];
        A[ix(t, c, occupancy)] = A[ix(t, c, occupancy)] + N;
        Sample the number of individuals n_next to be transitioned to each next compartment c_next
        C = sample_next_compartments(N, c);

        for (n_next, c_next) ∈ C do
            Sample n_next delays to be applied for this transition
            Δ = sample_delays(n_next, c, c_next);

            foreach δ_i ∈ Δ do
                Update the occupancy and inward-transitions slots to account for each transition

                A[ix(t + δ_i, c, occupancy)] = A[ix(t + δ_i, c, occupancy)] − 1;
                A[ix(t + δ_i, c_next, transitions)] = A[ix(t + δ_i, c_next, transitions)] + 1;
```

#### 3 Approximate Bayesian Computation example

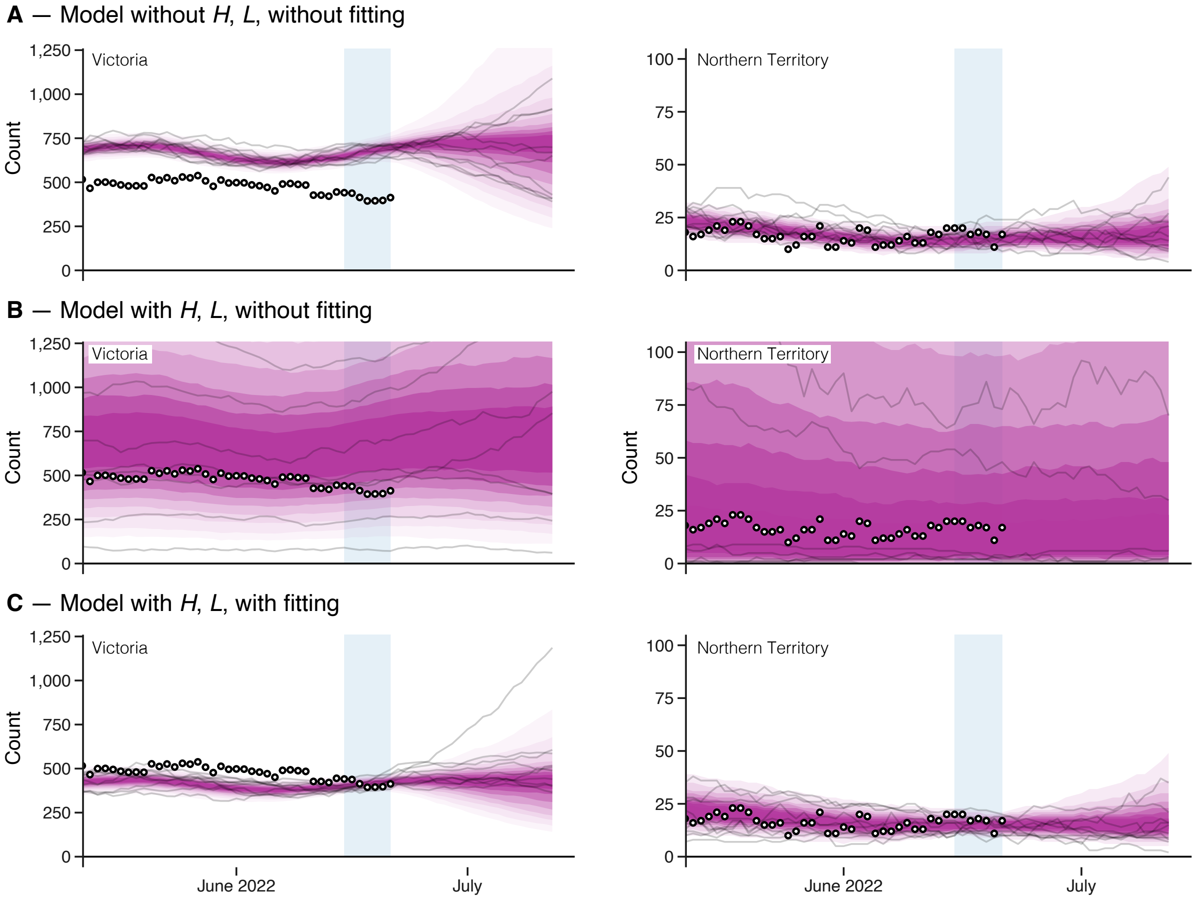


Supp. Methods, Figure 1: Model outputs for ward occupancy for Victoria (left) and the Northern Territory (right), produced retrospectively using data as at 2022-06-24 (forecast 15). Forecasts are displayed with intervals from 20% through to 90% in 10% increments are displayed in progressively lighter shading, with ten sample trajectories (in black) overlaid; reported ward occupancy counts are displayed as points. Light blue shading indicates the span of the 7-day calibration window. **A**: Model output for ward occupancy where there is no effect of the $H$ and $L$ parameters (i.e. $H = 0$, $L = 0$ across all simulations). Note that in this case, the distribution of trajectories for Victoria does not align with reported occupancy counts, whereas that for the Northern Territory does. **B**: Model output where $H$ and $L$ are sampled from prior distributions (see Methods). **C**: Final model outputs, with fitting, produced via rejection sampling from the outputs in **B**.

#### 4 Length of stay and transition probability estimates

Length of stay and transition probability estimates used in this study are presented below. Length of stay estimates are Gamma distribution parameters (shape, scale). Two sets of estimates were used throughout the study period. Table 1 describes estimates used in forecasts which were produced between March 18 2022 and 1 July 2022 (forecasts 1–16); these are the same estimates as presented in [1]. Table 2 describes estimates used in forecasts which were produced between 8 July 2022 and August 3 2022 (forecasts 17–21); these were produced using data as of 3 May 2022, across 5,693 patients who were admitted to hospitals in New South Wales between 1 March 2022 and 21 April 2022.

[1] Tobin, R. J., Wood, J. G., Jayasundara, D., Sara, G., Walker, C. R., Martin, G. E., McCaw, J. M., Shearer, F. M., & Price, D. J. (2023). Real-time analysis of hospital length of stay in a mixed SARS-CoV-2 Omicron and Delta epidemic in New South Wales, Australia. BMC Infectious Diseases, 23(1), 28.

| Pathway | Age | n | Scale | | Shape | | Probability | |
|---|---|---|---|---|---|---|---|---|
| ward-to-discharge | 0-39 | 4208 | 2.27 | [ 2.16, 2.39] | 0.95 | [0.92, 0.98] | 0.96 | [0.95, 0.96] |
| | 40-69 | 3014 | 4.98 | [ 4.68, 5.30] | 0.79 | [0.76, 0.83] | 0.87 | [0.86, 0.88] |
| | 70+ | 2701 | 7.32 | [ 6.71, 7.95] | 1.04 | [0.99, 1.09] | 0.78 | [0.76, 0.80] |
| ward-to-ICU | 0-39 | 162 | 1.48 | [ 1.09, 1.93] | 0.56 | [0.46, 0.67] | 0.04 | [0.03, 0.04] |
| | 40-69 | 402 | 3.81 | [ 3.12, 4.61] | 0.48 | [0.43, 0.53] | 0.11 | [0.10, 0.12] |
| | 70+ | 356 | 4.09 | [ 3.34, 4.95] | 0.50 | [0.45, 0.57] | 0.09 | [0.08, 0.10] |
| ward-to-death | all | 289 | 41.69 | [32.20, 53.31] | 0.82 | [0.72, 0.93] | 0.03 | [0.03, 0.03] |
| ICU-to-discharge | 0-69 | 62 | 10.71 | [ 5.86, 17.52] | 0.70 | [0.52, 0.92] | 0.13 | [0.10, 0.16] |
| | 70+ | 25 | 10.71 | [ 5.86, 17.52] | 0.70 | [0.46, 1.02] | 0.08 | [0.06, 0.11] |
| ICU-to-death | 0-69 | 48 | 20.60 | [13.91, 29.24] | 0.96 | [0.74, 1.23] | 0.13 | [0.10, 0.17] |
| | 70+ | 55 | 20.60 | [13.91, 29.24] | 0.86 | [0.67, 1.10] | 0.23 | [0.19, 0.28] |
| ICU-to-post-ICU | 0-69 | 391 | 4.15 | [ 3.58, 4.73] | 1.30 | [1.16, 1.46] | 0.74 | [0.70, 0.78] |
| | 70+ | 221 | 4.15 | [ 3.58, 4.73] | 1.37 | [1.21, 1.54] | 0.69 | [0.63, 0.74] |
| post-ICU-to-discharge | 0-39 | 110 | 4.34 | [ 3.58, 5.20] | 1.12 | [0.94, 1.34] | 0.97 | [0.90, 0.99] |
| | 40-69 | 209 | 4.34 | [ 3.58, 5.20] | 1.63 | [1.41, 1.88] | 0.93 | [0.85, 0.97] |
| | 70+ | 134 | 4.34 | [ 3.58, 5.20] | 2.00 | [1.70, 2.34] | 0.77 | [0.69, 0.84] |
| post-ICU-to-death | all | 24 | 15.97 | [ 1.44, 62.61] | 0.95 | [0.42, 1.84] | 0.05 | [0.05, 0.05] |

Table 1: Estimates used for forecasts 1–16 (18 March 2022 through to 1 July 2022). Length of stay and transition probability parameter estimate means and 95% confidence intervals, with sample size (n) and correlation (Cor.) between the natural logarithms of the estimated shape and scale parameters. Note that ward-to-death and post-ICU-to-death probability estimates were produced using the Aalen-Johansen non-parametric estimator and are depicted without uncertainty. Note that the ward-to-ICU probabilities were replaced with time-varying estimates in the forecasting model.

| Pathway | Age | n | Scale | | Shape | | Probability | |
|---|---|---|---|---|---|---|---|---|
| ward_to_discharge | 0-39 | 2181 | 2.32 | [ 2.16, 2.49] | 0.91 | [0.86, 0.96] | 0.95 | [0.94, 0.96] |
| | 40-69 | 1393 | 4.98 | [ 4.51, 5.45] | 0.74 | [0.69, 0.79] | 0.90 | [0.88, 0.91] |
| | 70+ | 1393 | 7.61 | [ 6.87, 8.43] | 0.98 | [0.91, 1.05] | 0.84 | [0.82, 0.86] |
| ward_to_ICU | 0-39 | 80 | 1.49 | [ 0.95, 2.10] | 0.54 | [0.41, 0.68] | 0.04 | [0.03, 0.04] |
| | 40-69 | 123 | 2.76 | [ 1.91, 3.86] | 0.44 | [0.36, 0.54] | 0.08 | [0.07, 0.09] |
| | 70+ | 130 | 3.58 | [ 2.53, 4.95] | 0.53 | [0.43, 0.64] | 0.07 | [0.06, 0.08] |
| ward_to_death | all | 93 | 90.65 | [53.24, 144.42] | 0.66 | [0.53, 0.81] | 0.02 | [0.02, 0.02] |
| ICU_to_discharge | 0-69 | 49 | 3.46 | [ 2.21, 5.19] | 1.20 | [0.84, 1.67] | 0.25 | [0.20, 0.32] |
| | 70+ | 15 | 3.46 | [ 2.21, 5.19] | 1.48 | [0.91, 2.29] | 0.12 | [0.07, 0.19] |
| ICU_to_death | 0-69 | 10 | 23.38 | [ 8.71, 49.40] | 0.79 | [0.40, 1.40] | 0.08 | [0.05, 0.13] |
| | 70+ | 17 | 23.38 | [ 8.71, 49.40] | 0.56 | [0.33, 0.88] | 0.17 | [0.11, 0.25] |
| ICU_to_postICU | 0-69 | 132 | 3.35 | [ 2.64, 4.19] | 1.30 | [1.07, 1.58] | 0.67 | [0.60, 0.73] |
| | 70+ | 90 | 3.35 | [ 2.64, 4.19] | 1.27 | [1.03, 1.58] | 0.71 | [0.62, 0.78] |
| postICU_to_discharge | 0-39 | 40 | 3.77 | [ 2.83, 4.96] | 1.35 | [1.03, 1.77] | 0.87 | [0.73, 0.95] |
| | 40-69 | 58 | 3.77 | [ 2.83, 4.96] | 1.67 | [1.29, 2.13] | 0.81 | [0.69, 0.89] |
| | 70+ | 66 | 3.77 | [ 2.83, 4.96] | 1.99 | [1.55, 2.49] | 0.84 | [0.73, 0.91] |
| postICU_to_death | all | 5 | 4.06 | [ 0.58, 14.65] | 1.49 | [0.37, 3.86] | 0.02 | [0.02, 0.02] |

Table 2: Estimates used for forecasts 17 to 21 (8 July 2022 to 3 August 2022). Length of stay and transition probability parameter estimate means and 95% confidence intervals, with sample size (n) and correlation (Cor.) between the natural logarithms of the estimated shape and scale parameters. Note that ward-to-death and post-ICU-to-death probability estimates were produced using the Aalen-Johansen non-parametric estimator and are depicted without uncertainty. Note that the ward-to-ICU probabilities were replaced with time-varying estimates in the forecasting model.

#### 5 Model changes throughout the study period

*Changes are dated by date of version control code commit.*

**2022-04-07**

We introduced specific adjustment factor prior distributions for the Northern Territory. Prior to this change, $\sigma^2_{\text{hosp}} = 0.8$ and $\sigma^2_{\text{los}} = 0.5$ was set across all jurisdictions. After this change, $\sigma^2_{\text{hosp}} = 2$ and $\sigma^2_{\text{los}} = 2$ was set for the Northern Territory, with priors for all other jurisdictions unchanged.

For calculation of the truncation-adjusted time-varying estimates, we increased the shape and scale of the Gamma distribution for the case-to-ward delay by a factor of 1.4.

**2022-04-27**

We reverted the delay distribution parameter changes which were made on 07-04-2022.

**2022-05-19**

The list of states excluded from using local data for the time-varying estimates was changed (from NT, SA, QLD and WA to NT, SA and QLD).

**2022-05-27**

The list of states excluded from using local data for the time-varying estimates was changed (from NT, SA, and QLD to NT, SA, QLD and VIC).

**2022-06-27**

We removed a modified on probability of hospitalisation which adjusted for cases which were detected by rapid antigen test (RAT) being missing from the New South Wales NNDSS dataset, as these cases were now present in the data. This adjustment had increased the probability of hospitalisation according to the number of missing RAT cases, with this missing count determined via a separately supplied publicly available case count.

The list of states excluded from using local data for the time-varying estimates was changed (from NT, SA, QLD and VIC to NT, SA and QLD).

**2022-07-06**

We introduced specific adjustment factor prior distributions for Queensland. After this change, $\sigma^2_{\text{hosp}} = 2$ and $\sigma^2_{\text{los}} = 2$ was set for both the Northern Territory and Queensland, with priors for all other jurisdictions unchanged.

**2022-07-08**

The list of states excluded from using local data for the time-varying estimates was changed (from NT, SA and QLD to NT, SA, QLD, TAS and VIC).

**2022-08-22**

The list of states excluded from using local data for the time-varying estimates was changed (from NT, SA, QLD, TAS and VIC to NT, SA and QLD).
