## Supplementary Forecast Performance for "A modular approach to forecasting COVID-19 hospital bed occupancy"

### Supplementary Materials — Forecast Performance

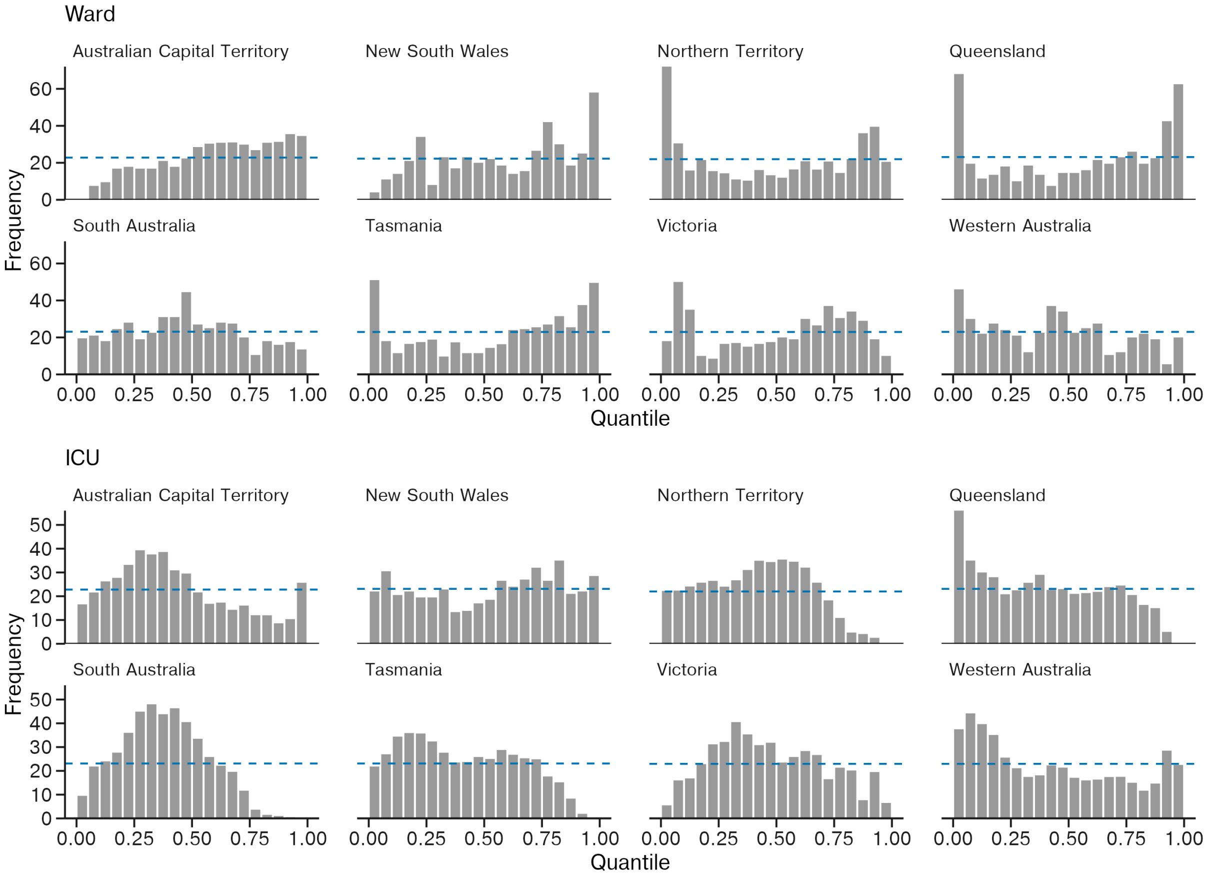


Supp. Performance, Figure 1: PIT (probability integral transform) plots for the ward and ICU forecasts produced across the study period. Each column represents the probability that an eventual reported observation fell within each predicted decile. Ideal calibration of the forecasts is achieved where the overall distribution is flat, such that all columns are at the height of the dashed horizontal line.

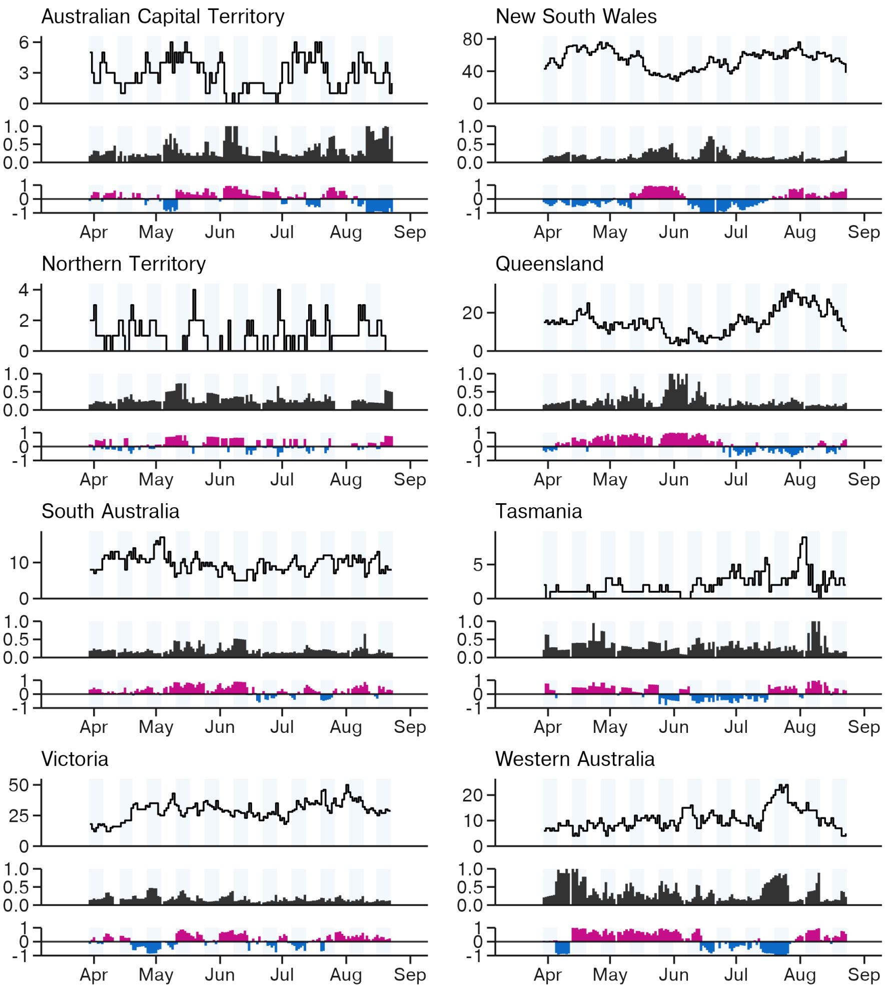


Supp. Performance, Figure 2: 15–21 day horizon performance of the ICU forecasts for the forecasts produced between March 18 and August 3 2022. The eventual true ICU occupancy count is displayed. Below, the CRPS and bias of the forecast is displayed, reflecting the performance of forecasted counts for that date. The CRPS displayed is calculated over log-transformed counts as described in the methods. Optimal forecasting performance is achieved where these values are closest to zero.

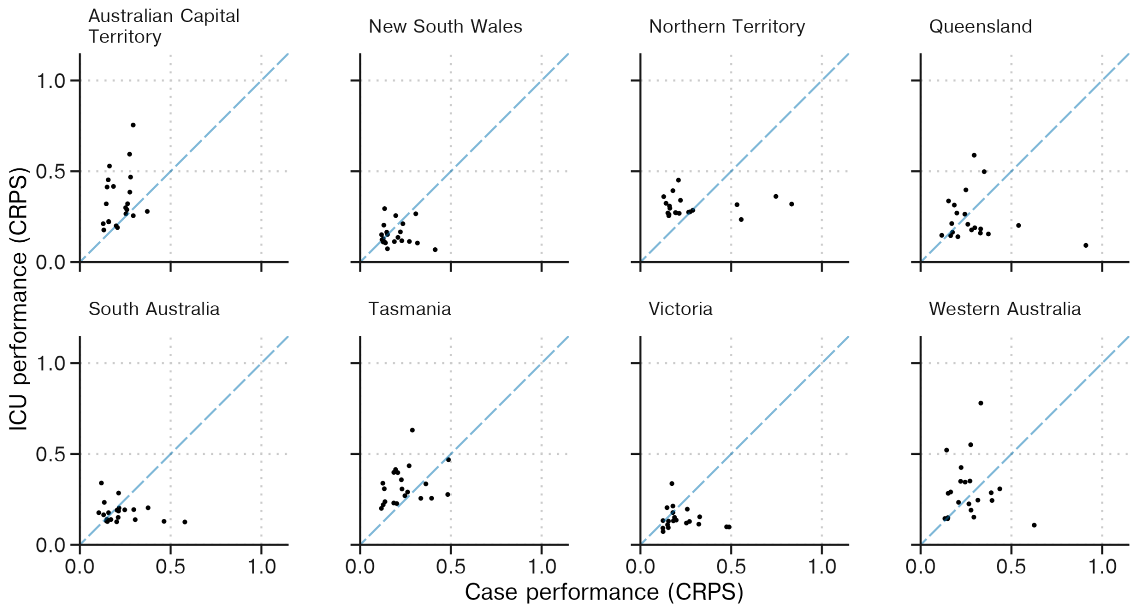


Supp. Performance, Figure 3: Performance of the ICU occupancy forecasts (y-axis) compared to the corresponding case forecast used to produce it (x-axis), where performance is measured using CRPS over log-transformed counts.

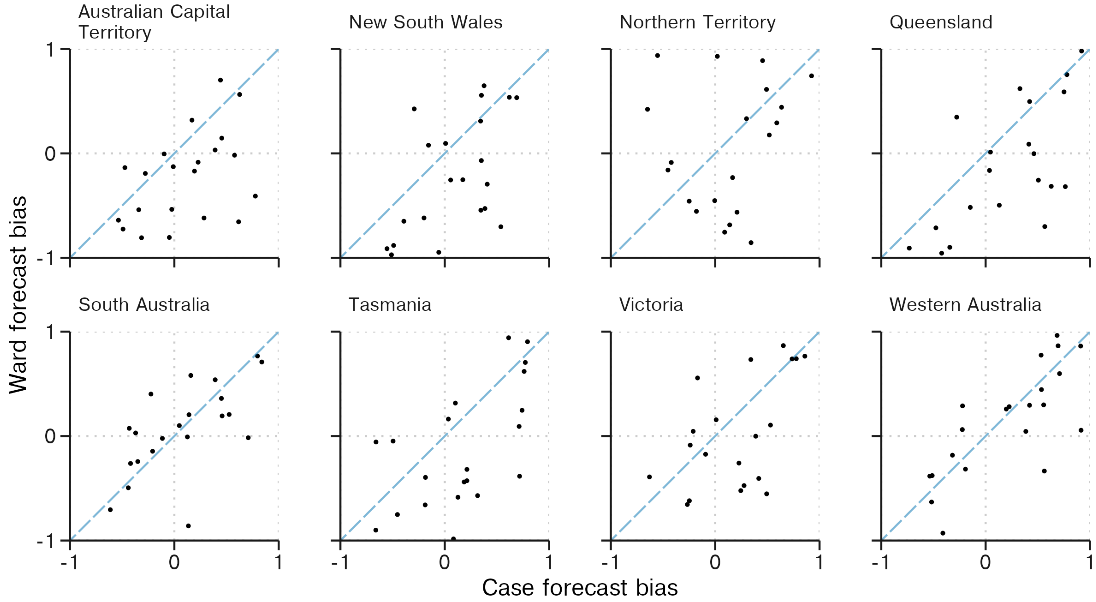


Supp. Performance, Figure 4: Performance of the ward occupancy forecasts (y-axis) compared to the corresponding case forecast used to produce it (x-axis), where performance is measured as bias.

#### Past forecasts – all states and territories

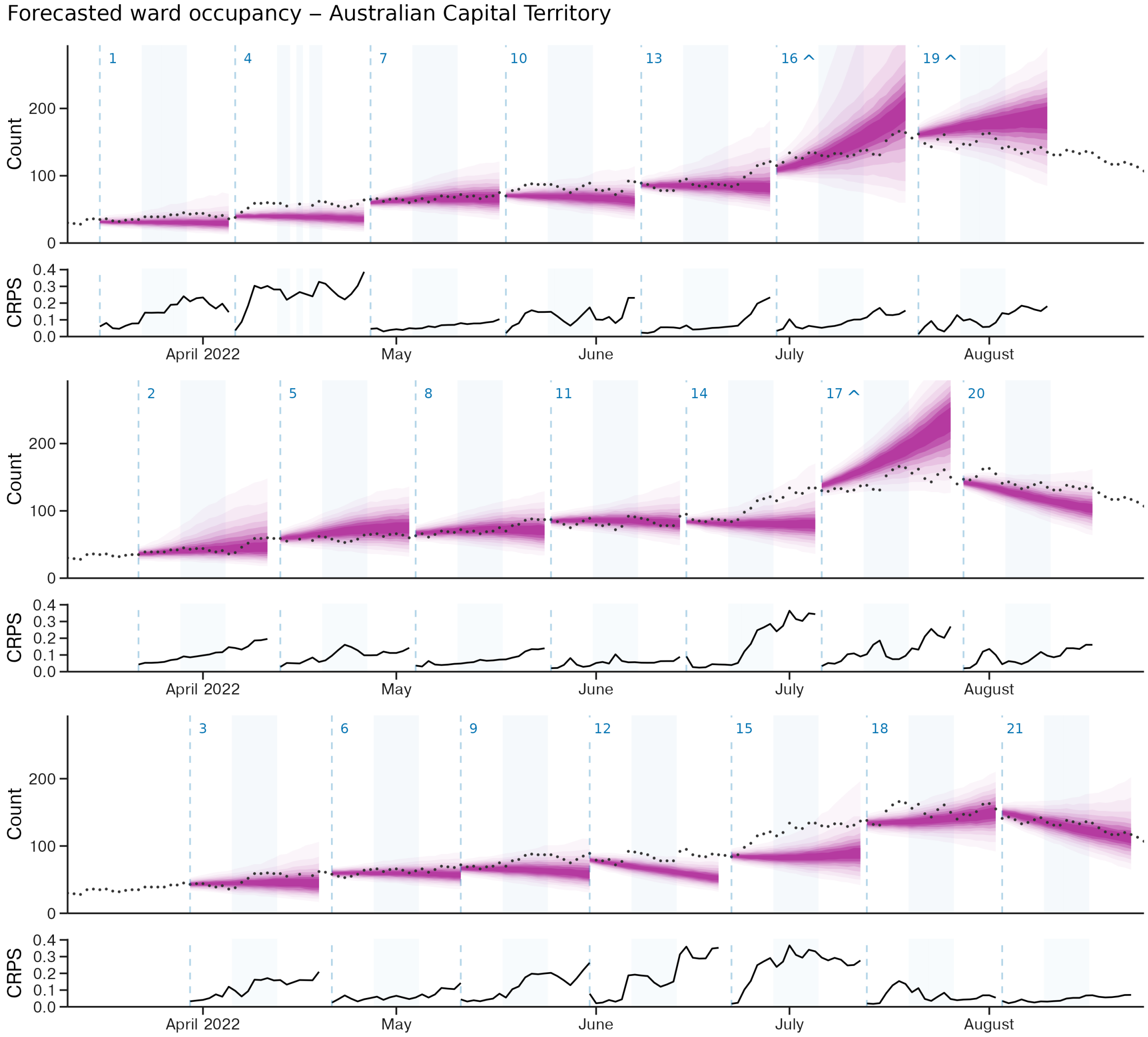


Supp. Performance, Figure 5: Forecasts for ward occupancy for the Australian Capital Territory produced between March 18 and August 3 2022. Credible intervals from 20% through to 90% in 10% increments are displayed in progressively lighter shading. CRPS values for log-transformed forecast predictions are displayed below each forecast. Forecast start dates are displayed as vertical dashed lines. Forecasts are plotted across three rows. The index of each forecast, 1 through 21, is displayed above each forecast start, with a ^ displayed where the upper quantiles of the forecast exceed the y-axis limits.

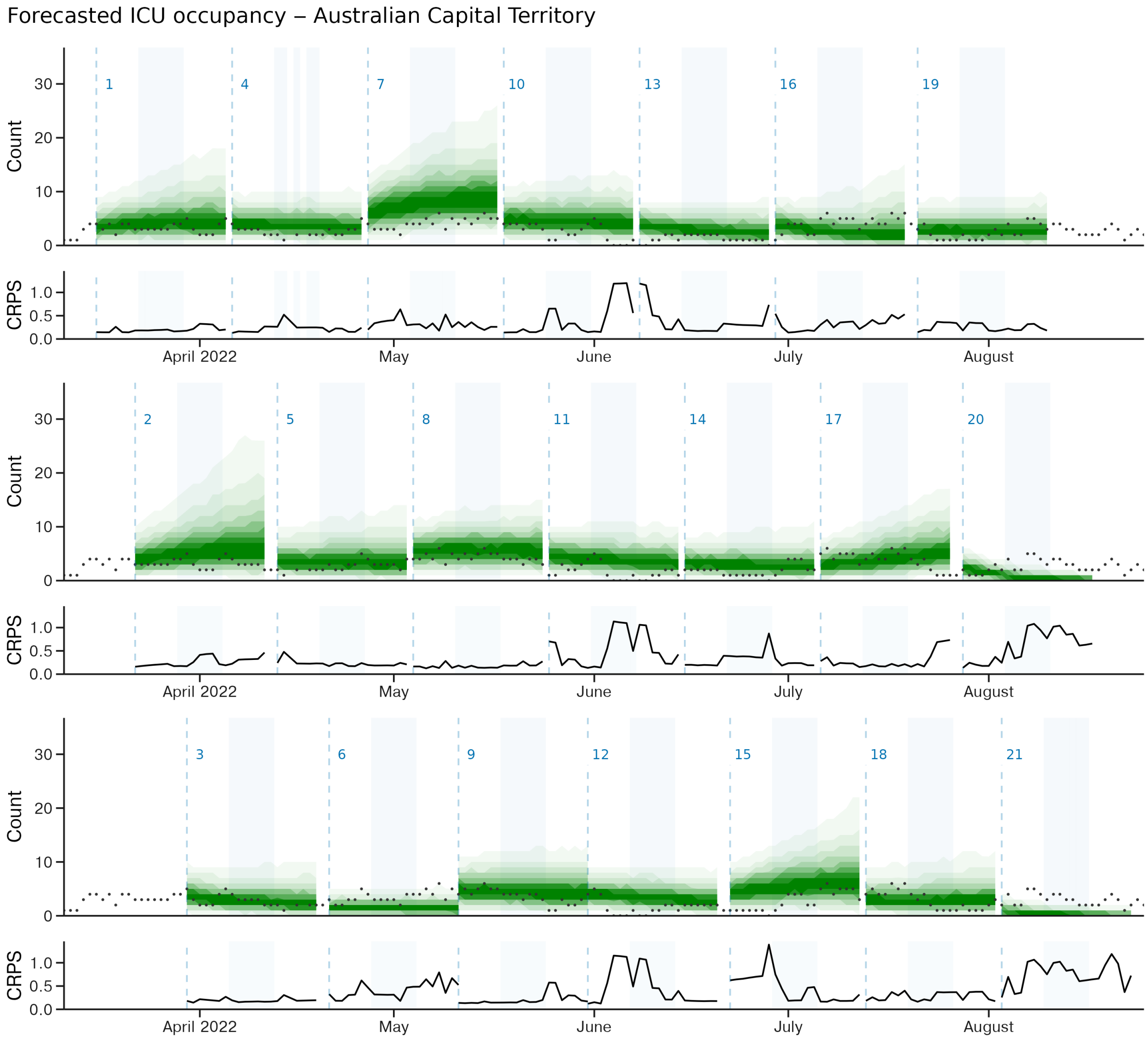


Supp. Performance, Figure 6: Forecasts for ICU occupancy for the Australian Capital Territory produced between March 18 and August 3 2022. Credible intervals from 20% through to 90% in 10% increments are displayed in progressively lighter shading. CRPS values for log-transformed forecast predictions are displayed below each forecast. Forecast start dates are displayed as vertical dashed lines. Forecasts are plotted across three rows. The index of each forecast, 1 through 21, is displayed above each forecast start, with a ^ displayed where the upper quantiles of the forecast exceed the y-axis limits.

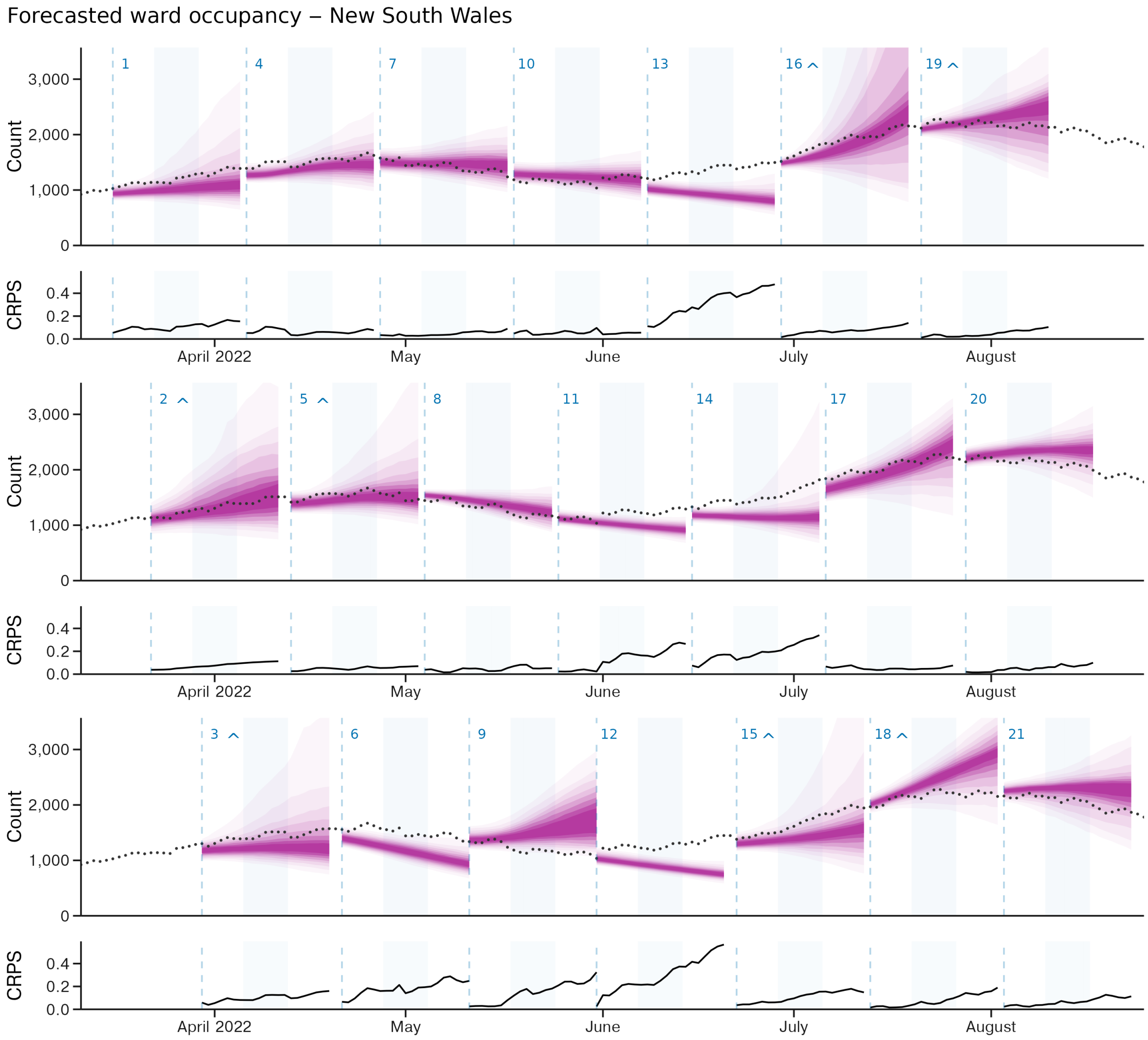


Supp. Performance, Figure 7: Forecasts for ward occupancy for the state of New South Wales produced between March 18 and August 3 2022. Credible intervals from 20% through to 90% in 10% increments are displayed in progressively lighter shading. CRPS values for log-transformed forecast predictions are displayed below each forecast. Forecast start dates are displayed as vertical dashed lines. Forecasts are plotted across three rows. The index of each forecast, 1 through 21, is displayed above each forecast start, with a ^ displayed where the upper quantiles of the forecast exceed the y-axis limits.

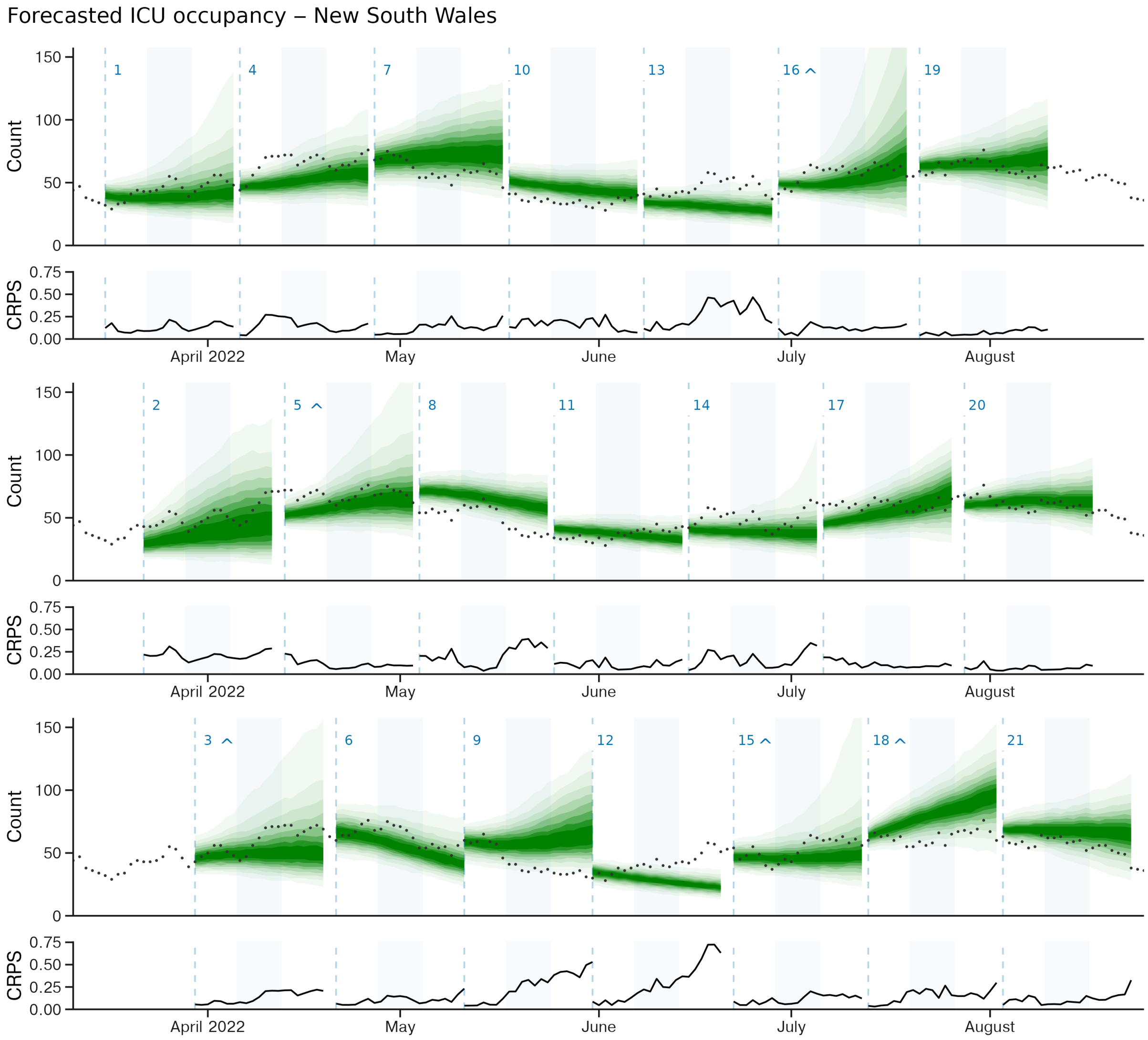


Supp. Performance, Figure 8: Forecasts for ICU occupancy for the state of New South Wales produced between March 18 and August 3 2022. Credible intervals from 20% through to 90% in 10% increments are displayed in progressively lighter shading. CRPS values for log-transformed forecast predictions are displayed below each forecast. Forecast start dates are displayed as vertical dashed lines. Forecasts are plotted across three rows. The index of each forecast, 1 through 21, is displayed above each forecast start, with a ^ displayed where the upper quantiles of the forecast exceed the y-axis limits.

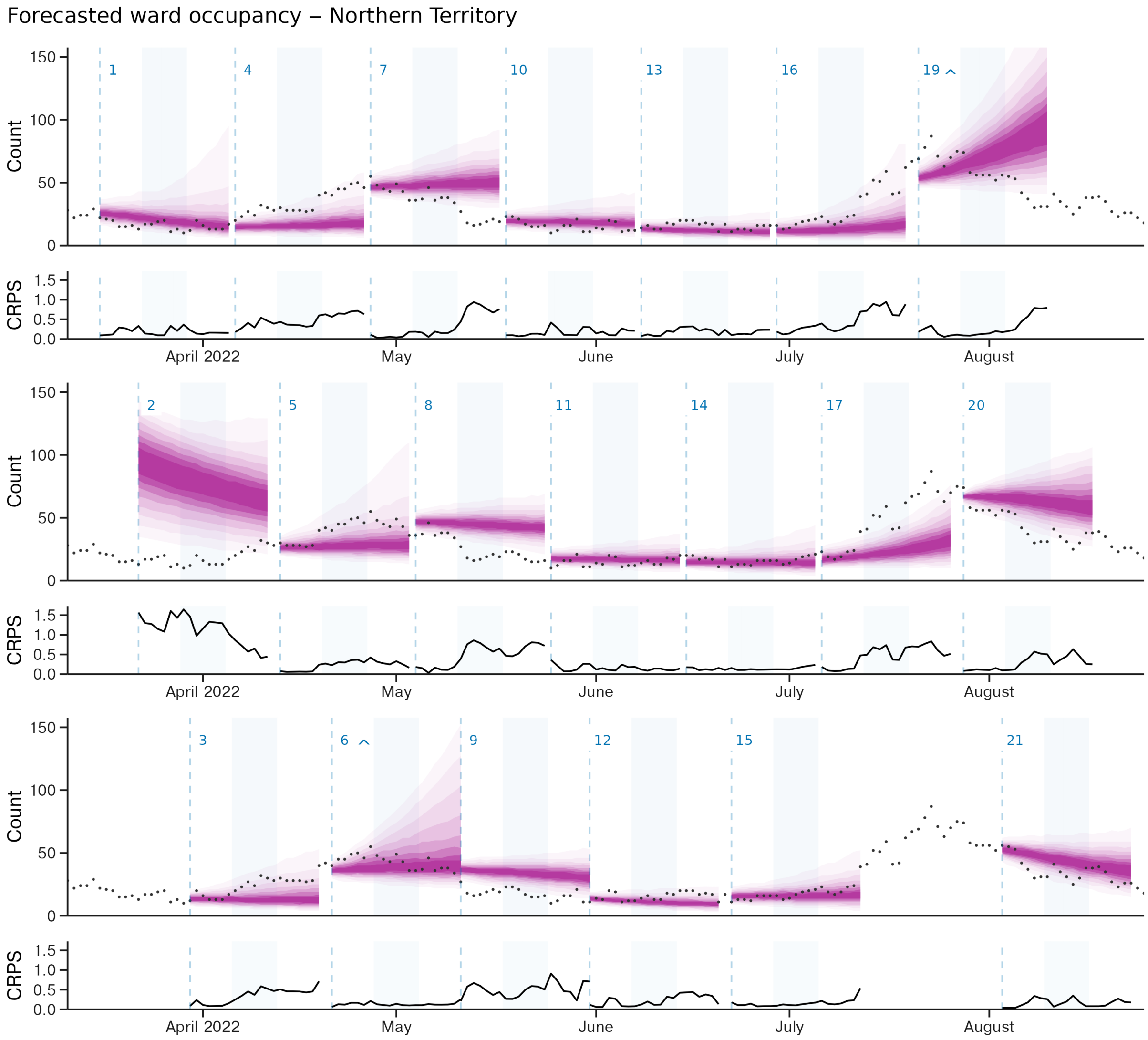


Supp. Performance, Figure 9: Forecasts for ward occupancy for the Northern Territory produced between March 18 and August 3 2022. Credible intervals from 20% through to 90% in 10% increments are displayed in progressively lighter shading. CRPS values for log-transformed forecast predictions are displayed below each forecast. Forecast start dates are displayed as vertical dashed lines. Forecasts are plotted across three rows. The index of each forecast, 1 through 21, is displayed above each forecast start, with a ^ displayed where the upper quantiles of the forecast exceed the y-axis limits. **Note forecast 2**: Poor fit due to failed ABC inference. This was addressed in later forecasts through modification of the $H$ and $L$ priors to increase prior predictive uncertainty (see Supplementary Methods). **Note forecast 18**: Not produced due to technical issues.

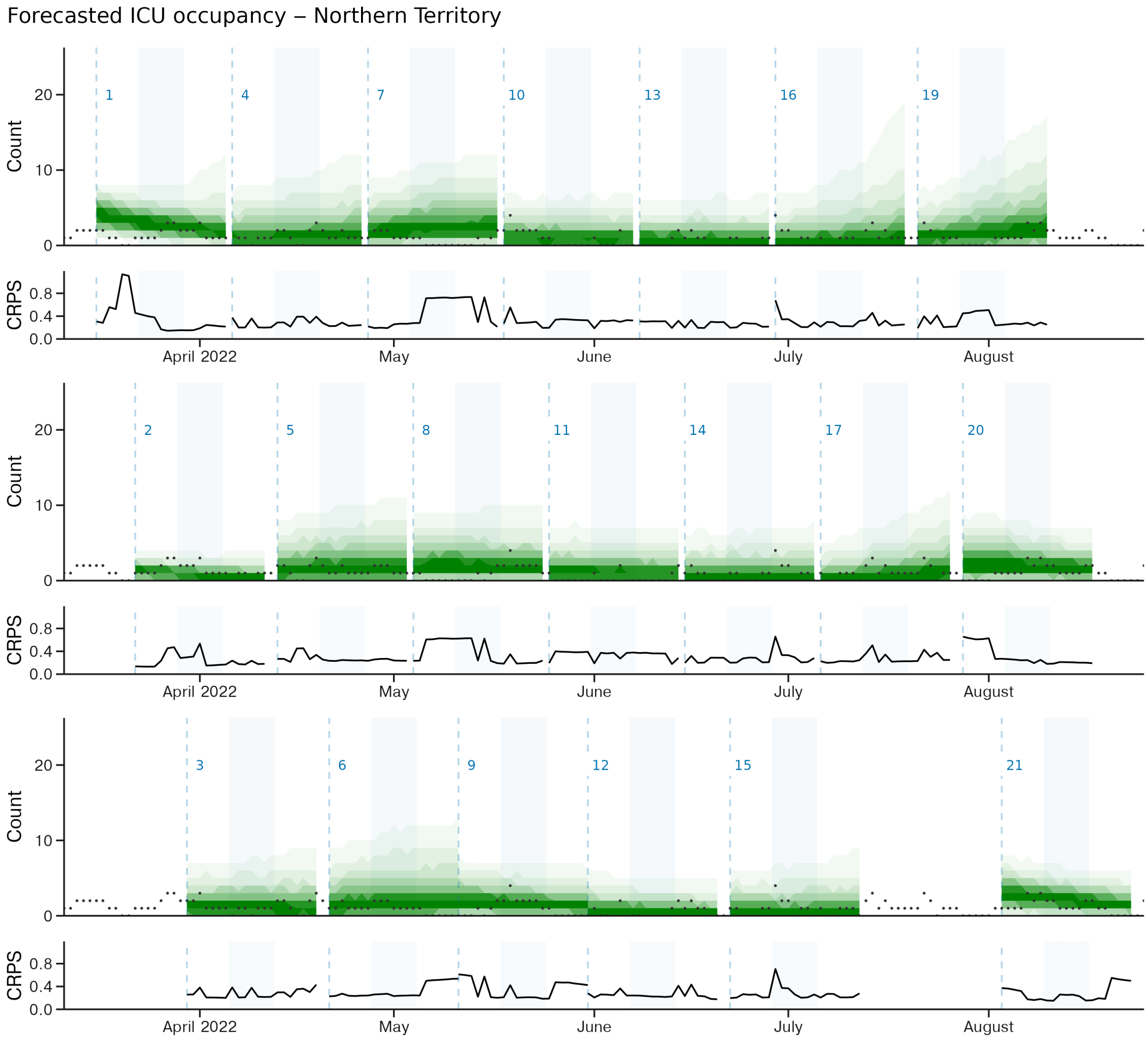


Supp. Performance, Figure 10: Forecasts for ICU occupancy for the Northern Territory produced between March 18 and August 3 2022. Credible intervals from 20% through to 90% in 10% increments are displayed in progressively lighter shading. CRPS values for log-transformed forecast predictions are displayed below each forecast. Forecast start dates are displayed as vertical dashed lines. Forecasts are plotted across three rows. The index of each forecast, 1 through 21, is displayed above each forecast start, with a ^ displayed where the upper quantiles of the forecast exceed the y-axis limits. **Note forecast 18**: Not produced due to technical issues.

Forecasted ward occupancy − Queensland

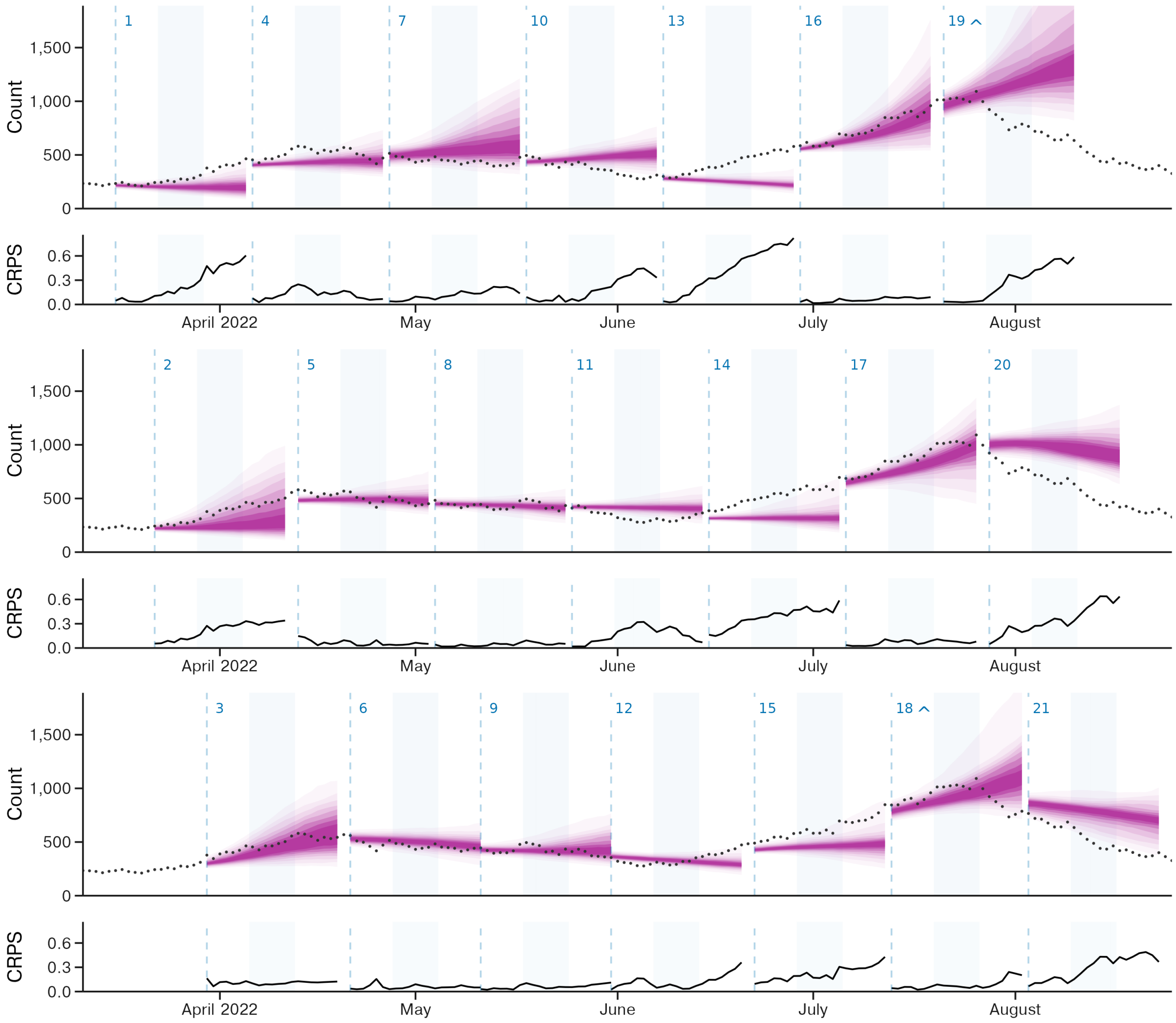


Supp. Performance, Figure 11: Forecasts for ward occupancy for the state of Queensland produced between March 18 and August 3 2022. Credible intervals from 20% through to 90% in 10% increments are displayed in progressively lighter shading. CRPS values for log-transformed forecast predictions are displayed below each forecast. Forecast start dates are displayed as vertical dashed lines. Forecasts are plotted across three rows. The index of each forecast, 1 through 21, is displayed above each forecast start, with a ^ displayed where the upper quantiles of the forecast exceed the y-axis limits.

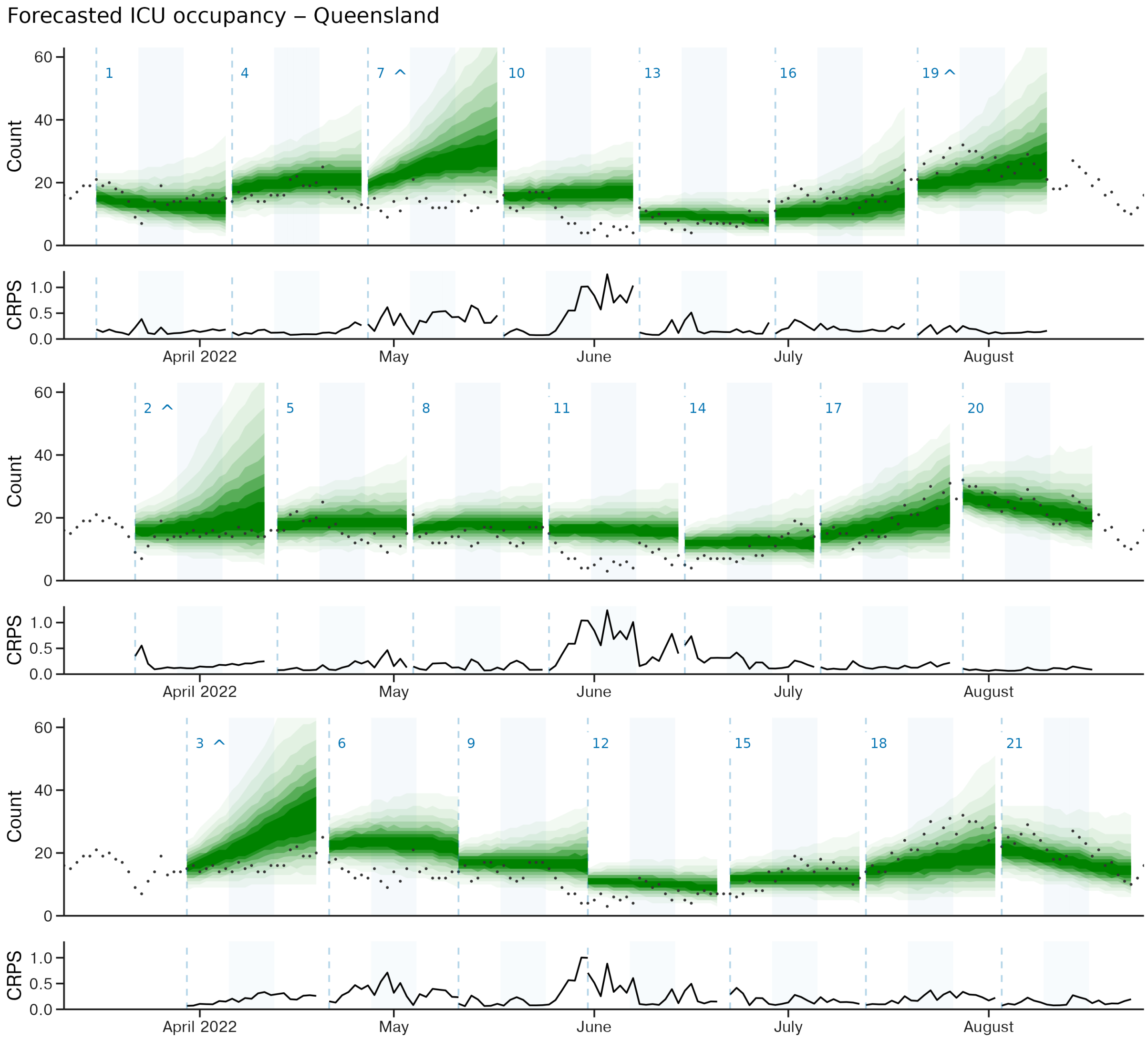


Supp. Performance, Figure 12: Forecasts for ICU occupancy for the state of Queensland produced between March 18 and August 3 2022. Credible intervals from 20% through to 90% in 10% increments are displayed in progressively lighter shading. CRPS values for log-transformed forecast predictions are displayed below each forecast. Forecast start dates are displayed as vertical dashed lines. Forecasts are plotted across three rows. The index of each forecast, 1 through 21, is displayed above each forecast start, with a ^ displayed where the upper quantiles of the forecast exceed the y-axis limits.

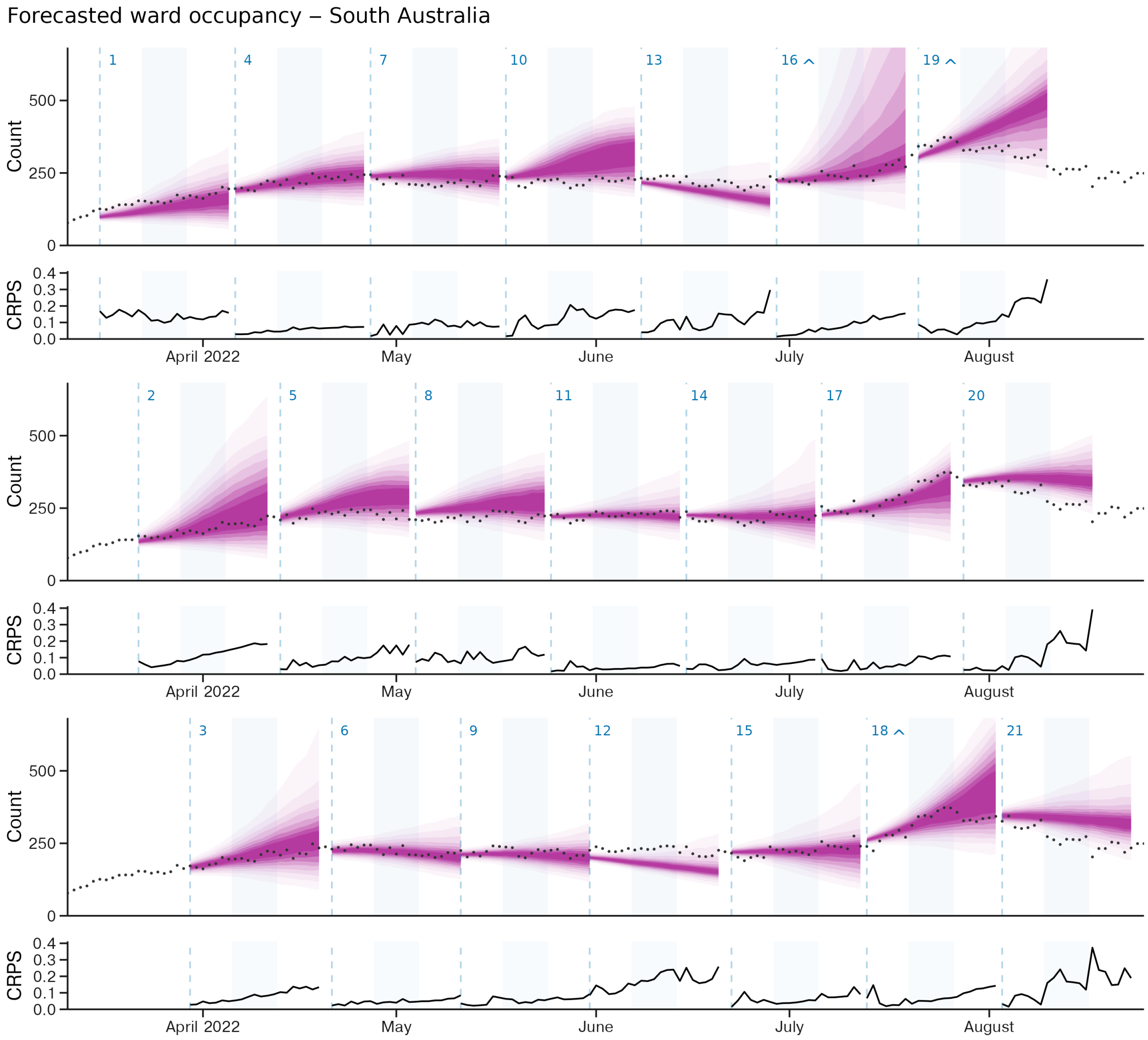


Supp. Performance, Figure 13: Forecasts for ward occupancy for the state of South Australia produced between March 18 and August 3 2022. Credible intervals from 20% through to 90% in 10% increments are displayed in progressively lighter shading. CRPS values for log-transformed forecast predictions are displayed below each forecast. Forecast start dates are displayed as vertical dashed lines. Forecasts are plotted across three rows. The index of each forecast, 1 through 21, is displayed above each forecast start, with a ^ displayed where the upper quantiles of the forecast exceed the y-axis limits.

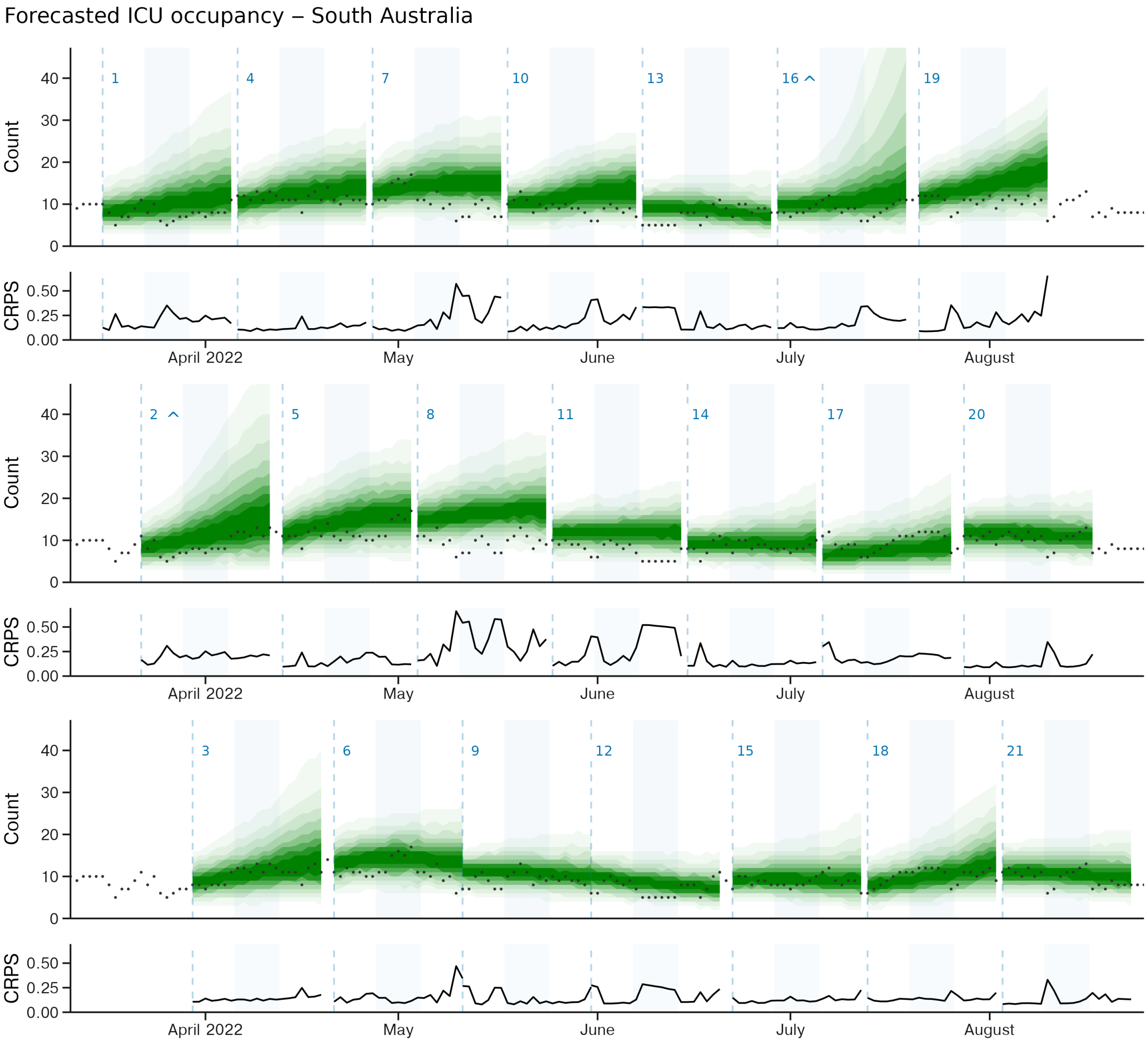


Supp. Performance, Figure 14: Forecasts for ICU occupancy for the state of South Australia produced between March 18 and August 3 2022. Credible intervals from 20% through to 90% in 10% increments are displayed in progressively lighter shading. CRPS values for log-transformed forecast predictions are displayed below each forecast. Forecast start dates are displayed as vertical dashed lines. Forecasts are plotted across three rows. The index of each forecast, 1 through 21, is displayed above each forecast start, with a ^ displayed where the upper quantiles of the forecast exceed the y-axis limits.

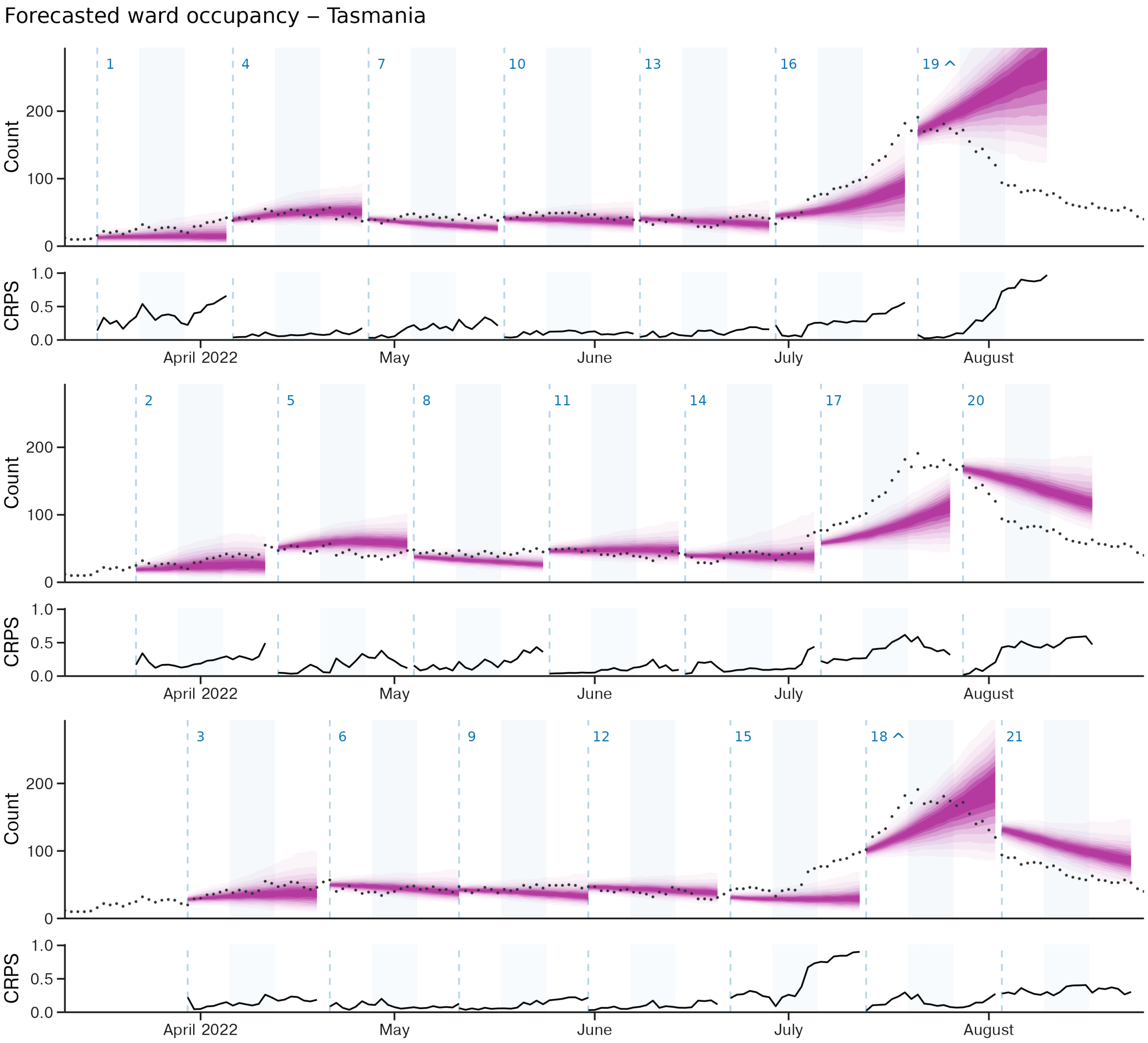


Supp. Performance, Figure 15: Forecasts for ward occupancy for the state of Tasmania produced between March 18 and August 3 2022. Credible intervals from 20% through to 90% in 10% increments are displayed in progressively lighter shading. CRPS values for log-transformed forecast predictions are displayed below each forecast. Forecast start dates are displayed as vertical dashed lines. Forecasts are plotted across three rows. The index of each forecast, 1 through 21, is displayed above each forecast start, with a ^ displayed where the upper quantiles of the forecast exceed the y-axis limits.

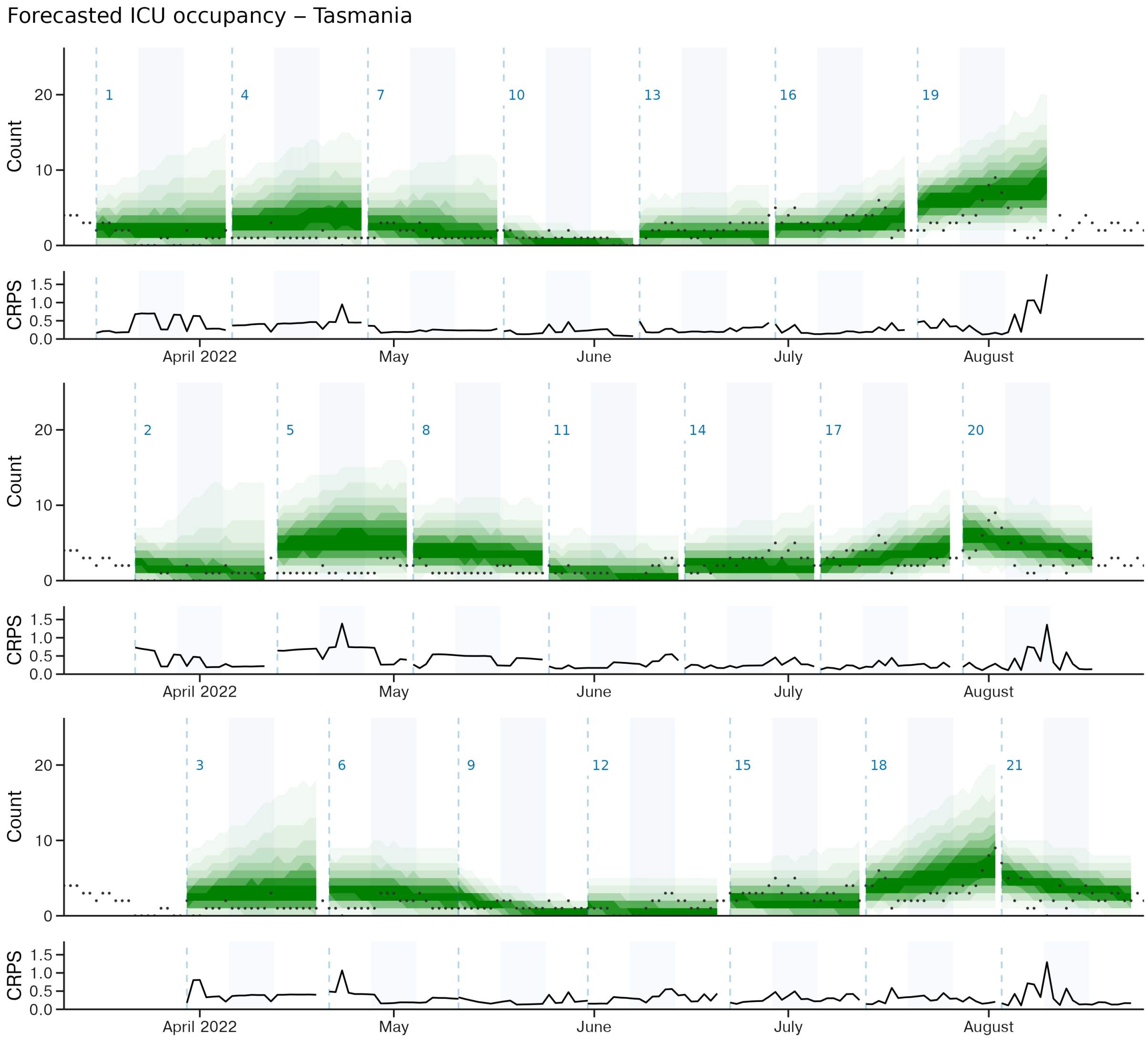


Supp. Performance, Figure 16: Forecasts for ICU occupancy for the state of Tasmania produced between March 18 and August 3 2022. Credible intervals from 20% through to 90% in 10% increments are displayed in progressively lighter shading. CRPS values for log-transformed forecast predictions are displayed below each forecast. Forecast start dates are displayed as vertical dashed lines. Forecasts are plotted across three rows. The index of each forecast, 1 through 21, is displayed above each forecast start, with a ^ displayed where the upper quantiles of the forecast exceed the y-axis limits.

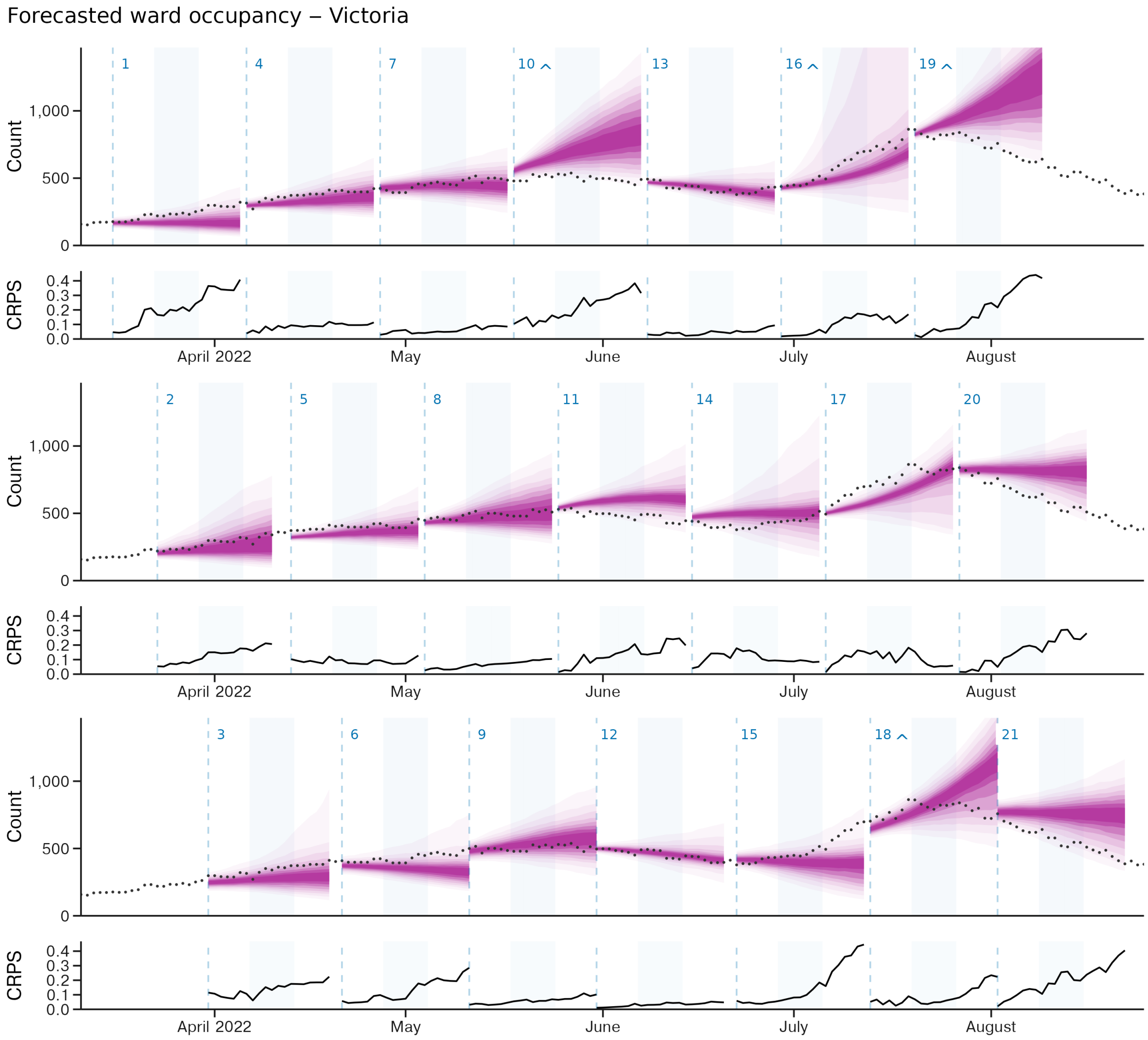


Supp. Performance, Figure 17: Forecasts for ward occupancy for the state of Victoria produced between March 18 and August 3 2022. Credible intervals from 20% through to 90% in 10% increments are displayed in progressively lighter shading. CRPS values for log-transformed forecast predictions are displayed below each forecast. Forecast start dates are displayed as vertical dashed lines. Forecasts are plotted across three rows. The index of each forecast, 1 through 21, is displayed above each forecast start, with a ^ displayed where the upper quantiles of the forecast exceed the y-axis limits.

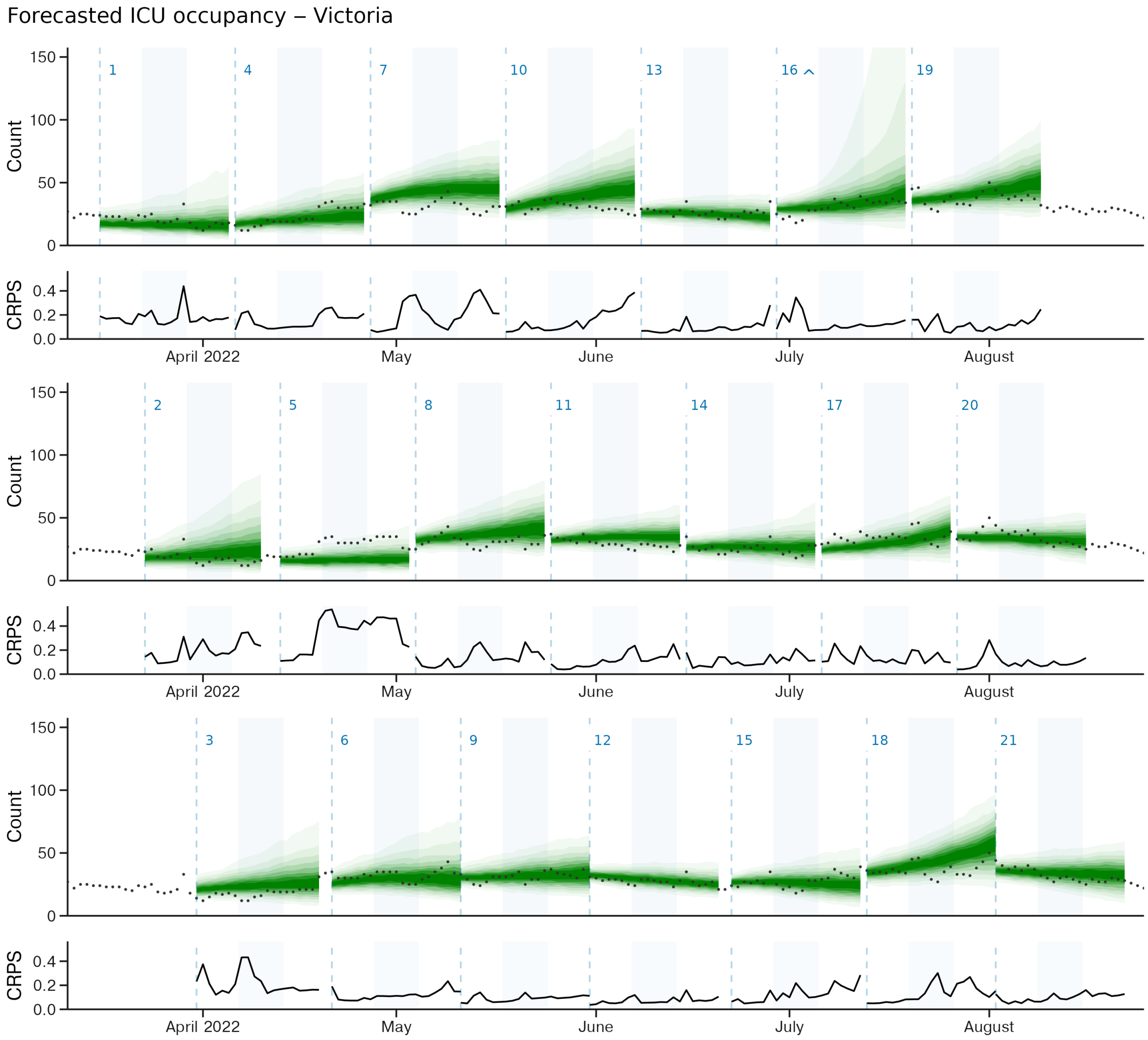


Supp. Performance, Figure 18: Forecasts for ICU occupancy for the state of Victoria produced between March 18 and August 3 2022. Credible intervals from 20% through to 90% in 10% increments are displayed in progressively lighter shading. CRPS values for log-transformed forecast predictions are displayed below each forecast. Forecast start dates are displayed as vertical dashed lines. Forecasts are plotted across three rows. The index of each forecast, 1 through 21, is displayed above each forecast start, with a ^ displayed where the upper quantiles of the forecast exceed the y-axis limits.

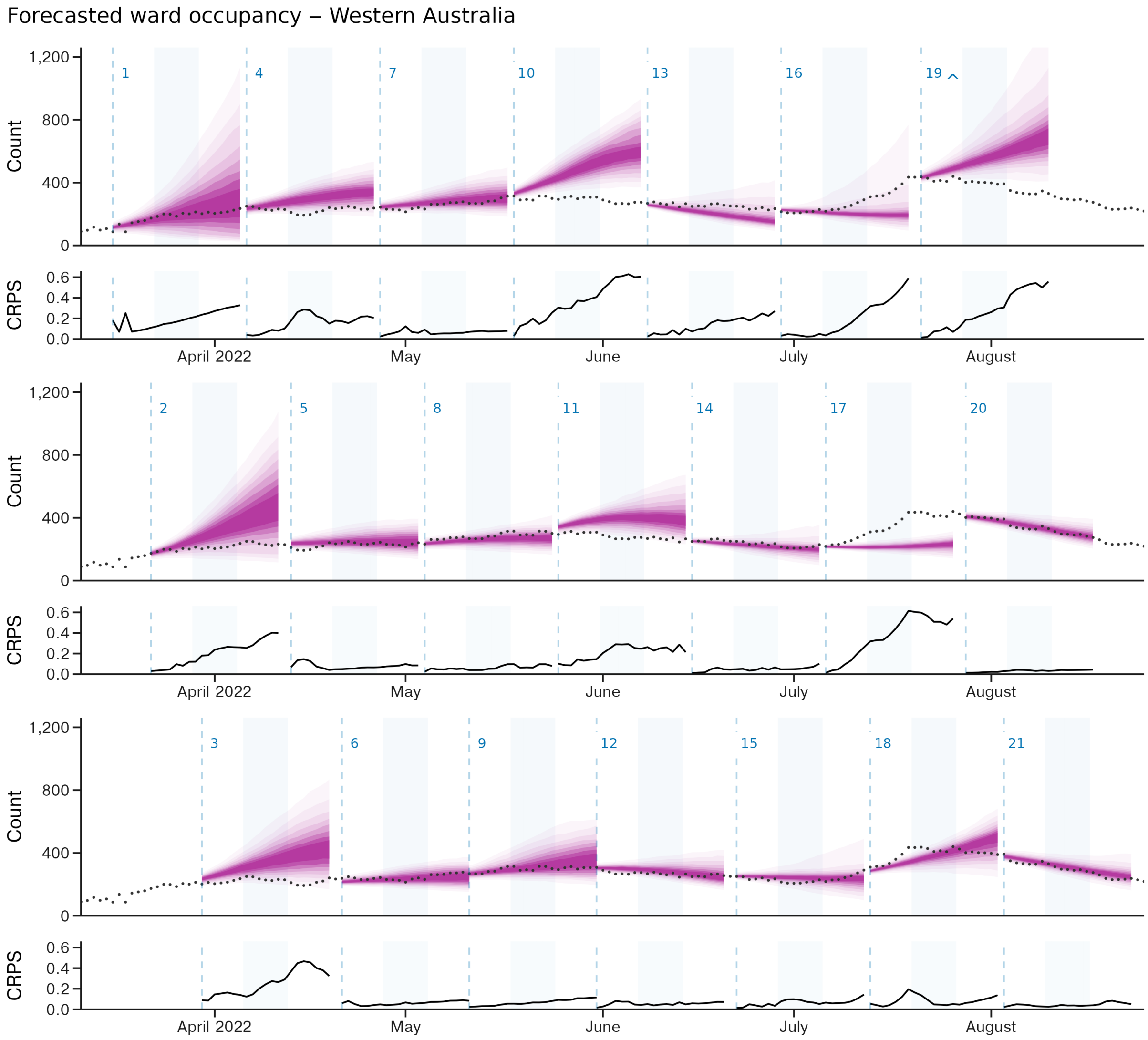


Supp. Performance, Figure 19: Forecasts for ward occupancy for the state of Western Australia produced between March 18 and August 3 2022. Credible intervals from 20% through to 90% in 10% increments are displayed in progressively lighter shading. CRPS values for log-transformed forecast predictions are displayed below each forecast. Forecast start dates are displayed as vertical dashed lines. Forecasts are plotted across three rows. The index of each forecast, 1 through 21, is displayed above each forecast start, with a ^ displayed where the upper quantiles of the forecast exceed the y-axis limits.

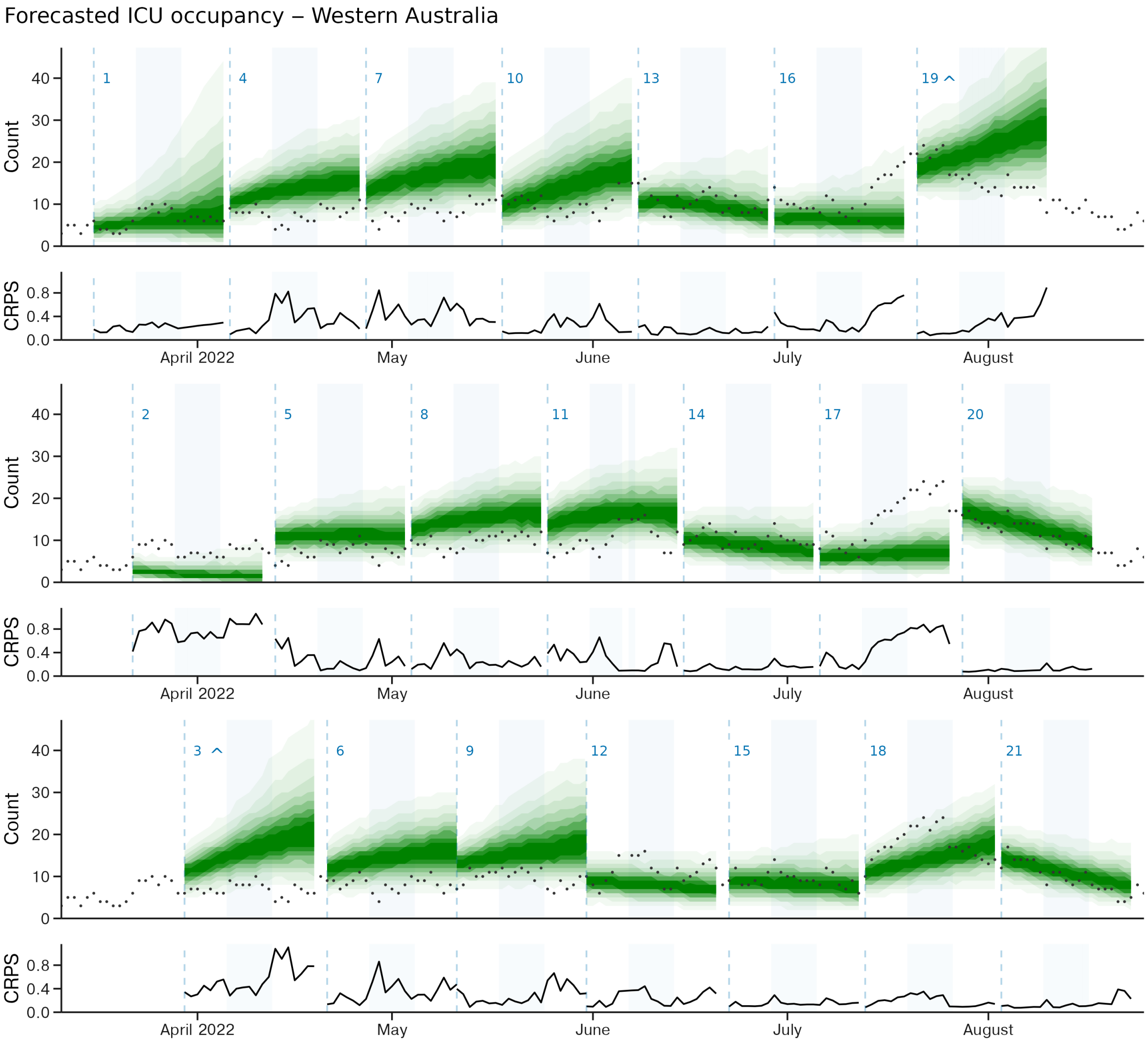


Supp. Performance, Figure 20: Forecasts for ICU occupancy for the state of Western Australia produced between March 18 and August 3 2022. Credible intervals from 20% through to 90% in 10% increments are displayed in progressively lighter shading. CRPS values for log-transformed forecast predictions are displayed below each forecast. Forecast start dates are displayed as vertical dashed lines. Forecasts are plotted across three rows. The index of each forecast, 1 through 21, is displayed above each forecast start, with a ^ displayed where the upper quantiles of the forecast exceed the y-axis limits.
